# Asymptomatic and symptomatic SARS-CoV-2 infections elicit polyfunctional antibodies

**DOI:** 10.1101/2020.11.12.20230508

**Authors:** Jérémy Dufloo, Ludivine Grzelak, Isabelle Staropoli, Yoann Madec, Laura Tondeur, François Anna, Stéphane Pelleau, Aurélie Wiedemann, Cyril Planchais, Julian Buchrieser, Rémy Robinot, Marie-Noëlle Ungeheuer, Hugo Mouquet, Pierre Charneau, Michael White, Yves Lévy, Bruno Hoen, Arnaud Fontanet, Olivier Schwartz, Timothée Bruel

## Abstract

A large proportion of SARS-CoV-2 infected individuals remains asymptomatic. Little is known about the extent and quality of their antiviral humoral response. Here, we analyzed antibody functions in 52 asymptomatic infected individuals, 119 mild and 21 hospitalized COVID-19 patients. We measured anti-Spike antibody levels with the S-Flow assay and mapped SARS-CoV-2 Spike- and N-targeted regions by Luminex. Neutralization, complement deposition and Antibody-Dependent Cellular Cytotoxicity (ADCC) were evaluated using replication-competent SARS-CoV-2 or reporter cell systems. We show that COVID-19 sera mediate complement deposition and kill infected cells by ADCC. Sera from asymptomatic individuals neutralize the virus, activate ADCC and trigger complement deposition. Antibody levels and activities are slightly lower in asymptomatic individuals. The different functions of the antibodies are correlated, independently of disease severity. Longitudinal samplings show that antibody functions follow similar kinetics of induction and contraction, with minor variations. Overall, asymptomatic SARS-CoV-2 infection elicits polyfunctional antibodies neutralizing the virus and targeting infected cells.

- Sera from convalescent COVID-19 patients activate the complement and kill infected cells by ADCC.
- Asymptomatic and symptomatic SARS-CoV-2-infected individuals harbor polyfunctional antibodies.
- Antibody levels and functions are slightly lower in asymptomatic individuals
- The different antiviral activities of anti-Spike antibodies are correlated regardless of disease severity.
- Functions of anti-Spike antibodies have similar kinetics of induction and contraction.

## Introduction

The Severe Acute Respiratory Syndrome Coronavirus 2 (SARS-CoV-2) emerged in 2019 and became pandemic in 2020 (Wu et al., 2020; Zhou et al., 2020a). SARS-CoV-2 is responsible for the Coronavirus disease 2019 (COVID-19) (Gorbalenya et al., 2020). As of Nov 4, 2020, almost 50 million individuals were infected and 1.2 million have died of COVID-19. The rapid spread of the virus has overwhelmed health care organization in many areas. In the absence of prophylactic or therapeutic strategies, governments used non-pharmaceutical measures to decrease viral transmission, such as school closures, mobility restrictions, curfews and global shutdown (Flaxman et al., 2020). As restrictive policies were relaxed, many countries experienced new epidemic waves, demonstrating the necessity for population immunity, triggered either by infection or vaccination, to limit SARS-CoV-2 circulation. Therefore, the immune response induced by SARS-CoV-2 is under intense investigation, aiming at informing vaccine design, identifying correlates of protection and determining the duration of protective immunity.

The outcome of SARS-CoV-2 infection is highly variable, ranging from asymptomatic disease to life-threatening acute respiratory distress syndrome (ARDS) (Guan et al., 2020; Huang et al., 2020). About half of infected individuals remain asymptomatic (Lavezzo et al., 2020; Sakurai et al., 2020). Males, the elderly, and people suffering from diabetes, obesity and cardiovascular diseases have an increased risk of admission to intensive care unit (ICU) and death (Cummings et al., 2020; Huang et al., 2020). Severe COVID-19 is due to immunological dysfunctions, including impaired type I interferon response (Bastard et al., 2020; Blanco-Melo et al., 2020; Hadjadj et al., 2020; Zhang et al., 2020) increased inflammation (Giamarellos-Bourboulis et al., 2020; Lucas et al., 2020; Zhou et al., 2020b), complement activation (Carvelli et al., 2020) and endothelial stress (Varga et al., 2020). Because of this over-activation of the immune system and prolonged antigenic exposure, survivors of severe COVID-19 display a strong immune memory to the virus, as determined by antibody titers and CD4^+^ T cell re-stimulation (Grzelak et al., 2020; Peng et al., 2020). Mild and asymptomatic infections induce seroconversion and production of neutralizing antibody (Fafi-Kremer et al., 2020; Long et al., 2020), but titers are lower in asymptomatic individuals (Long et al., 2020). Whether such responses are protective is unknown. A deeper understanding of the immune response after asymptomatic SARS-CoV-2 infection is needed.

The Spike protein of SARS-CoV-2 is responsible for viral entry by interacting with the angiotensin-converting enzyme 2 (ACE2) receptor (Hoffmann et al., 2020). It is a class I fusion protein, which requires proteolytic cleavage for activation and fusogenicity. The two subunits S1 and S2 assemble into a trimer of heterodimers to form the mature Spike (Wrapp et al., 2020). The S1 subunit samples “closed” and “open” conformations, the latter exposing the receptor binding domain (RBD), the main target of neutralizing antibodies (Barnes et al., 2020; Robbiani et al., 2020; Rogers et al., 2020; Wec et al., 2020). The Spike protein is exposed at the surface of viral particles and infected cells (Buchrieser et al., 2020), making them sensitive to antibody targeting. Antiviral activities of antibodies are not restricted to neutralization of viral particles. Infected cells covered by antibodies can be eliminated through various mechanisms, including antibody-dependent cellular cytotoxicity (ADCC) and complement-dependent cytotoxicity (CDC) (Bredow et al., 2016; Bruel et al., 2016; Dai et al., 2017; Dufloo et al., 2019; Richard et al., 2018). Those activities rely on the fragment crystallizable (Fc) domain of antibodies and cognate Fc Receptors (FcR)(Bournazos and Ravetch, 2017). Fc-effector functions are necessary for optimal efficacy of antibodies in both therapeutic and prophylactic settings (Bournazos et al., 2014; DiLillo et al., 2016; Gunn et al., 2018). Little is known about Fc-effector functions in SARS-CoV-2 infection. Antibodies in sera from critical COVID-19 patients form immune complexes that can trigger NK cell activation and complement deposition (Atyeo et al., 2020). However, whether such polyfunctional antibodies are induced during asymptomatic COVID-19 disease and whether they are able to eliminate infected cells remains unknown.

Here, we established cellular assays to evaluate ADCC and complement activities of sera from COVID-19 patients with different disease severities. We combined these assays with measurements of antibody titers and neutralization. Altogether, our results indicate that SARS-CoV-2 asymptomatic infection induces a polyfunctional antibody response. The evaluation of Fc-effector antibody functions is important to understand immune responses in COVID-19 patients and vaccinees.

## Results

### Sera from COVID-19 patients activate the complement

We first determined whether antibodies from COVID-19 patients may activate the complement after binding to SARS-CoV-2-infected cells. We used as target cells the lung epithelial cell line A549 expressing ACE2 (A549-ACE2 cells) (Buchrieser et al., 2020). Cells were infected with SARS-CoV-2 at a multiplicity of infection (MOI) of 1. After 24h, cells were incubated with heat-inactivated (HI) serum from either pre-pandemic or COVID-19 individuals as a source of antibodies (dilution 1:100). We selected a panel of 11 sera among SARS-CoV-2 seropositive individuals identified in two previous sero-epidemiological studies (Fontanet et al., 2020a, 2020b) **(Table 1)**. Sera sampled before 2019 (pre-pandemic samples) were used as controls (n=12). The source of complement was normal human serum (NHS) (dilution 1:2). Heat-inactivated serum (HIHS) was used as control **(Figure 1A)**. After 4h of incubation, cells were stained with anti-C3b/iC3b and anti-Spike antibodies to examine complement deposition and SARS-CoV-2 infection, respectively. C3b/iC3b deposition (later referred to as C3 deposition) was measured in Spike+ cells. As shown in one representative experiment **(Figure 1B)**, we did not observe C3 deposition in the absence of antibodies nor with a pre-pandemic serum. In contrast, with a COVID-19 serum, C3 deposition occurred on 79% of infected cells **(Figure 1B)**. To calculate a cutoff of positivity, we determined the mean signal of the 12 pre-pandemic sera and added 3 standard deviations (S.D.) **(Figure 1C and Supplementary Figure 1A)**. All but 1 COVID-19 serum displayed C3b deposition above this cutoff (p<0.0001; Mann-Whitney test) **(Figure 1C and Supplementary Figure 1A)**. No C3 deposition was observed with HIHS **(Figure 1A and Supplementary Figure 1B)**. Despite C3 deposition, we did not observe killing (complement-dependent cytotoxicity, CDC) of infected cells **(Figure 1D and Supplementary Figure 1C)**. We also used U2OS-ACE2-GFP-Split cells (also termed S-Fuse cells) (Buchrieser et al., 2020). This reporter cell line allows quantification and live imaging of SARS-CoV-2 replication by measuring the formation of GFP+ syncytia upon infection. Similarly, we detected complement deposition, but no CDC of S-Fuse infected cells (not shown).

**Table 1.**
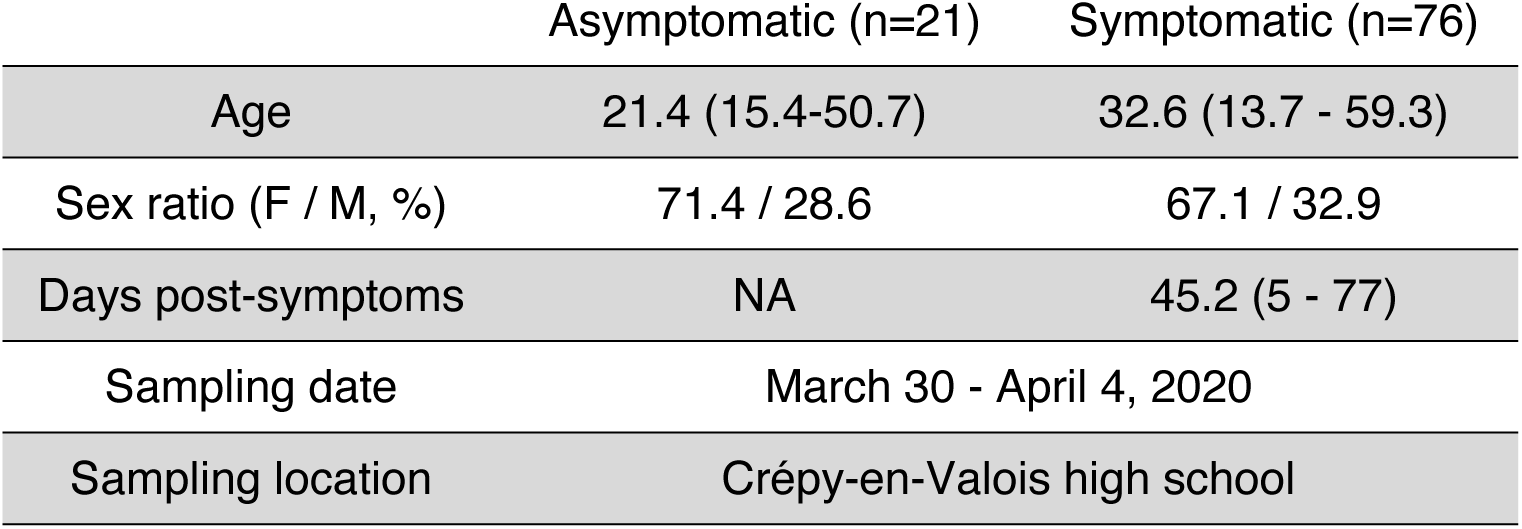
Characteristics of the first group of asymptomatic and symptomatic individuals.

|  | Asymptomatic (n=21) | Symptomatic (n=76) |
| --- | --- | --- |
| Age | 21.4 (15.4-50.7) | 32.6 (13.7 - 59.3) |
| Sex ratio (F / M, %) | 71.4 / 28.6 | 67.1 / 32.9 |
| Days post-symptoms | NA | 45.2 (5 - 77) |
| Sampling date | March 30 - April 4, 2020 |  |
| Sampling location | Crépy-en-Valois high school |  |

**Figure 1.**
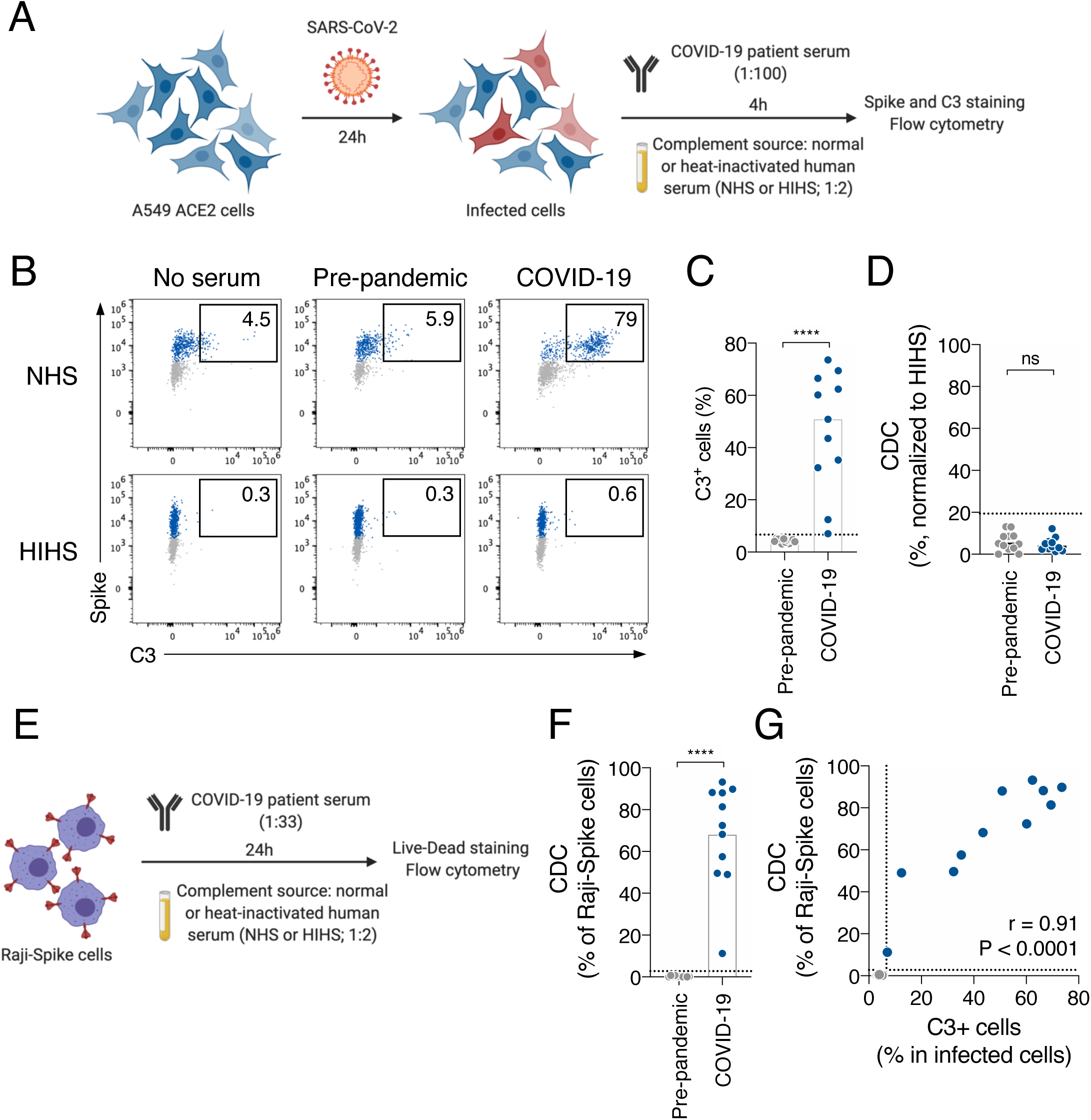
COVID-19 sera activate the complement. **A**. Schematic of the complement activation test on infected cells. A549-ACE2 cells are infected at a multiplicity of infection (MOI) of 1. 24 hours later, serum from pre-pandemic or COVID-19 patient is added (dilution 1:100) as a source of antibodies. Normal human serum (NHS) from a healthy donor is used as a source of complement and heat-inactivated human serum (HIHS) as control. 4 hours later, cells are stained with an anti-Spike and an anti-C3b/iC3b antibody. Complement deposition on infected cells is measured by flow cytometry. Complement-dependent cytotoxicity (CDC) induced by a serum is also measured as the relative disappearance of infected cells compared to the “HIHS” condition. **B**. Complement deposition on infected cells was measured after culture with or without a control or a COVID-19 serum in presence of NHS or HIHS. Results from a representative experiment are shown. Percentages indicate the proportion of C3+ cells among infected (Spike+) cells. **C**. Complement deposition with pre-pandemic (n=12) and COVID-19 patients’ (n=11) sera. The percentage of C3+ cells among infected cells is represented. Each dot represents the mean of 3 independent experiments for one serum donor. **D**. Complement-dependent cytotoxicity (CDC) of infected A549-ACE2 cells was calculated as the relative disappearance of Spike+ cells in the NHS compared to HIHS condition, with pre-pandemic (n=12) and COVID-19 patients’ (n=11) sera. Each dot represents the mean of 3 independent experiments for one serum donor. **E**. Schematic of the complement activation test on Raji-Spike cells. Raji-Spike cells are incubated with serum (heat-inactivated) from a pre-pandemic or a COVID-19 patient (dilution 1:100) and normal human serum (NHS) or heat-inactivated human serum (HIHS) as control (dilution 1:2). Complement-dependent cytotoxicity (CDC) induced by a serum is calculated as the relative cell death compared to the “no antibody” condition. **F**. Raji-Spike cells were cultured with sera from pre-pandemic individuals (n=11) or COVID-19 patients (n=11) and serum from a healthy individual as a source of complement. CDC was measured as the relative cell death compared to the no antibody condition. Each dot represents a different serum. **G**. Correlation of the C3 deposition on A549-ACE2 infected cells and CDC of Raji-Spike cells induced by sera from pre-pandemic individuals (grey, n=12) and COVID-19 patients (blue, n=11). In each graph, the dotted line corresponds to the positivity threshold calculated with pre-pandemic sera. In C, D and F, the bar indicates the mean and a Mann-Whitney test was performed (ns. = not significant, ****p<0.0001). In G, a Spearman correlation test was performed and the correlation r and p-value are indicated.

To evaluate the contribution of anti-Spike antibodies on complement deposition, we engineered Raji cells expressing the Spike protein. We chose Raji cells because they lack CD59, a potent CDC inhibitor (Dufloo et al., 2019). Raji-Spike cells were incubated with heat inactivated serum from either pre-pandemic or convalescent COVID-19 individuals (dilution 1:33), and normal human serum (NHS) as a source of complement (dilution 1:2). After 24h, cell death was assessed using a viability dye **(Figure 1E)**. COVID-19 sera displayed CDC activity above the threshold determined with the pre-pandemic sera (p<0.0001; Mann-Whitney test) **(Figure 1F)**. The CDC activity on Raji-Spike cells positively correlated with C3 deposition on A549-ACE2 infected cells (r=0.91, p<0.0001, Spearman correlation) **(Figure 1G)**.

### Sera from COVID-19 patients trigger Antibody-Dependent Cellular Cytotoxicity (ADCC)

We then assessed the ADCC activity of sera from COVID-19 patients. To visualize ADCC, we used SARS-CoV-2 infected S-Fuse cells as targets (Buchrieser et al., 2020). S-Fuse cells were infected for 18h with SARS-CoV-2 at a MOI of 0.1 and incubated with heterologous primary NK cells (ratio 1:1) in the presence or absence of serum (dilution 1:100). After 4h of co-culture, the area of infected (GFP+) cells was quantified using an automated microscope **(Figure 2A)**. GFP+ syncytia formed after infection were readily visible **(Figure 2B)**. Adding NK cells (ratio 1:1) decreased the GFP area, to a similar extent in “no serum” and “pre-pandemic serum” conditions, likely due to basal cytotoxic activity of NK cells against infected cells independently of any antibody **(Figure 2B)**. The GFP signal almost completely disappeared in the presence of a convalescent COVID-19 serum **(Figure 2B)**, demonstrating an induction of ADCC by NK cells. Of note, A549-ACE2 infected cells were not sensitive to ADCC (not shown). We thus used S-Fuse cells to interrogate our panel of COVID-19 sera and pre-pandemic samples using six donors of NK cells **(Figure 2C)**. We calculated the extent of GFP elimination triggered by each serum in comparison to the condition with NK cells and no serum. The 6 NK donors were active in this test, with variations in their efficacy **(Supplementary Figure 2)**. The mean value of pre-epidemic samples was used to determine a threshold of positivity (mean + 3 standard deviations (S.D)). Using this method, 10 out of the 11 tested COVID-19 sera displayed ADCC activity (p<0.0001; Mann-Whitney test) **(Figure 2C)**.

**Figure 2.**
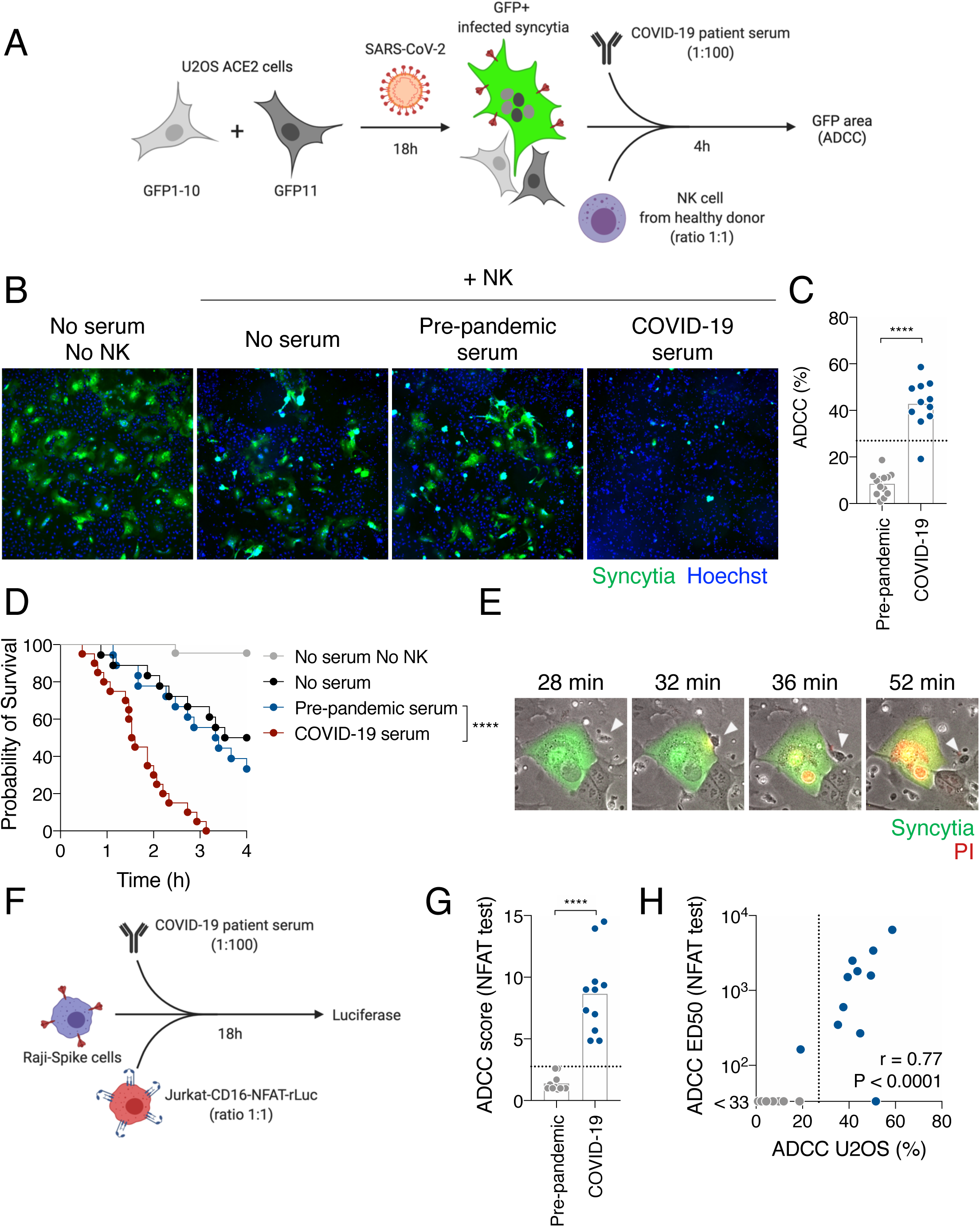
COVID-19 sera trigger antibody-dependent cellular cytotoxity by NK cells. **A**. Schematic of the ADCC test on infected cells. U2OS-ACE2-GFPsplit cells are infected with SARS-CoV-2 at a multiplicity of infection (MOI) of 1 for 18h. Cell-cell fusion between infected and neighboring cells reconstitute GFP. Serum from a pre-pandemic or COVID-19 individual (dilution 1:100) and NK cells from a healthy donor (at a 1:1 ratio) are added for 4h before quantification of the GFP area by high content imaging. ADCC is measured as the relative disappearance of the GFP area compared to the “no serum” condition. **B**. Representative images of co-culture of infected-U2OS-ACE2-GFPsplit cells with or without NK cells, and with or without serum from a pre-pandemic individual or a COVID-19 patient. Infected cells created GFP+ syncytia (green), allowing to track and quantity SARS-CoV-2 infection. Nuclei were stained with Hoechst (blue). **C**. Quantification of the antibody-dependent cellular cytotoxicity (ADCC) triggered by pre-pandemic (n=13) or COVID-19 (n=11) sera. Each dot represents the mean ADCC obtained with 6 donors of NK cells for one serum donor. **D**. The survival of infected U2OS-ACE2-GFPsplit cells was followed over 4 hours by live imaging without NK cells (grey, n=22) or with NK cells and no serum (black, n=18), a pre-pandemic serum (blue, n=18) or a COVID-19 serum (red, n=20). Results from two independent experiments are represented. **E**. Time-lapse of the ADCC of an infected cell by a NK cell in the presence of a COVID-19 serum. The syncytia appear in green (GFP). PI was added to monitor cell death and appears in red. The NK cell that comes and contacts the infected cell is indicated with a white arrowhead. Time post addition of NK cells and serum is indicated. **F**. Schematic of the CD16 activation test. Raji-Spike are co-cultured with Jurkat-CD16-NFAT-rLuc (ratio 1:1) in presence or absence of a pre-pandemic or COVID-19 serum (dilution 1:100). After 18 hours, luciferase expression (which occurs upon CD16 stimulation) is quantified and ADCC induced by a serum is calculated as the fold induction of luciferase expression compared to the “no serum” condition. **G**. Raji-Spike cells were co-cultured with Jurkat-CD16-NFAT-rLuc in presence of pre-pandemic (n=11) or COVID-19 (n=11) sera for 18h. The ADCC score was measured as the fold increase of Luciferase expression over the “no serum” condition. **H**. Correlation of the ADCC and the NFAT test score ED50 with sera from pre-pandemic individuals (grey, n=12) and COVID-19 patients (blue, n=11). In each graph, the dotted line corresponds to the positivity threshold calculated with pre-pandemic sera. In C and G, the bar indicates the mean and a Mann-Whitney test was performed (****p<0.0001). In D, a log-rank (Mantel-Cox) test was performed (****p<0.0001). In F, a Spearman correlation test was performed, and the correlation r and p-value are indicated.

We then performed video-microscopy experiments to determine whether the decrease of GFP was due to a killing of infected cells. SARS-CoV-2-infected S-Fuse cells were cultivated with NK cells and COVID-19 or pre-pandemic sera. Propidium iodide (PI) was added to the culture medium to monitor cell death. An image was recorded every 4 min with an automated time-lapse microscope. The infected GFP+ cells were tracked, and their viability was determined **(Figure 2D)**. After 4h of co-culture, 100% of infected cells were killed when NK cells and COVID-19 serum were present **(Figure 2D)**. NK cells alone or with a pre-pandemic serum reduced S-Fuse viability to a lower extent (p<0.0001; log-rang Mantel-Cox test) **(Figure 2D)**. Analysis of the movies revealed that NK cells established contacts with infected cells prior to death **(Figure 2E and video 1)**. These results show that sera from COVID-19 patients kill SARS-CoV-2-infected cell by ADCC.

We also used Raji-Spike cells and Jurkat-CD16-NFAT-rLuc reporter cells as a simple assay to evaluate ADCC of COVID-19 sera. This system measures the capacity of a serum to activate NFAT through CD16 (the pathway initiating ADCC in NK cells) in the presence of antigen-expressing target cells **(Figure 2F)**. The convalescent sera activated CD16 above the threshold calculated with the pre-pandemic samples (p<0.0001; Mann-Whitney test) **(Figure 2G)**. The extent of CD16 activation correlated to ADCC activity against S-Fuse infected cells, suggesting that anti-Spike antibodies contribute to the killing of infected cells (r=0.77, p<0.0001; Spearman correlation) **(Figure 2H)**.

Overall, these data show that sera from SARS-CoV-2 infected individuals harbor polyfunctional antibodies triggering complement deposition and ADCC. NK cells rapidly eliminate infected cells in the presence of specific antibodies, whereas complement activation is non-lytic, at least in the cell lines tested here.

### Antibody responses in asymptomatic and mild COVID-19 individuals

We then took advantage of two sero-epidemiological cohort studies conducted in a high school and in a primary school, respectively (Fontanet et al., 2020a, 2020b), to determine whether sera of COVID-19 asymptomatic and symptomatic individuals differ in their ability to trigger ADCC and complement deposition. In the first cohort, we identified a group of 21 seropositive individuals, who did not report any symptoms (asymptomatic individuals; AS) (**Table 1)**. As a control, we randomly selected seropositive individuals who reported at least one symptom (symptomatic individuals, S; n=76 among 137). These individuals suffered from a mild disease, as none required external oxygen supplementation nor hospitalization. The symptomatic group reported their first symptoms on average 45 days prior to blood sampling **(Table 1)**. AS were significantly younger than S individuals (21.4 vs 32.6 years old; **Table 1)**.

With the sera from these 98 individuals, we first measured the presence of antibodies binding the Spike, at a 1:300 dilution **(Figure 3A)**. We adapted our S-Flow assay, which allows sensitive and quantitative assessment of anti-Spike IgG by flow cytometry (Grzelak et al., 2020), to also measure IgA and IgM **(Figure 3A)**. Consistently with our prior identification as being seropositive, AS and S individuals harbored IgG antibodies, as determined by a percentage of Spike+ cells above the cutoff of 20% **(Figure 3A)**. The frequency of IgA^+^ and IgM^+^ cells tended to be lower in AS individuals with no significant differences **(Figure 3A)**. We did not observe differences by analyzing the intensity of binding for individuals positive for IgG, IgA and IgM **(Supplementary Figure 3A)**. We then measured IgG binding titers by serial dilutions. Titration curves of AS were consistently below S individuals **(Figure 3B)**. Effective dilution 50% titers (ED50) were significantly lower in AS (p=0.018; Mann-Whitney test) **(Figure 3B)**. As the Spike protein contains multiple epitopes (Barnes et al., 2020), we used Luminex to map anti-S1, anti-S2, anti-RBD and anti-Spike IgGs **(Figure 3C)**. We also included the N protein **(Figure 3C)**, as well as antigens from hCoV-229E and hCoV-NL63, and from Adenovirus 40, Influenza A, Mumps and Rubella viruses, to assess pre-existing immunity to human seasonal coronaviruses and other viruses **(Supplementary Figure 3)**. The overall response to S1, S2, RBD, Spike and N antigens tended to be lower in AS than in S albeit not significant **(Figure 3C and Supplementary Figure 3B)**. We did not observe differences between AS and S in the IgG response to hCoV-229E and NL63 **(Supplementary Figure 3C)**. The response to the antigens from the other viruses was similar between the two groups, except for Influenza A, likely reflecting the age difference (p=0.001; Mann-Whitney test) **(Supplementary Figure 3D)**. Altogether, these data show that the intensity of the antibody response specific to SARS-CoV-2 is slightly lower in asymptomatic than in mildly symptomatic individuals.

**Figure 3.**
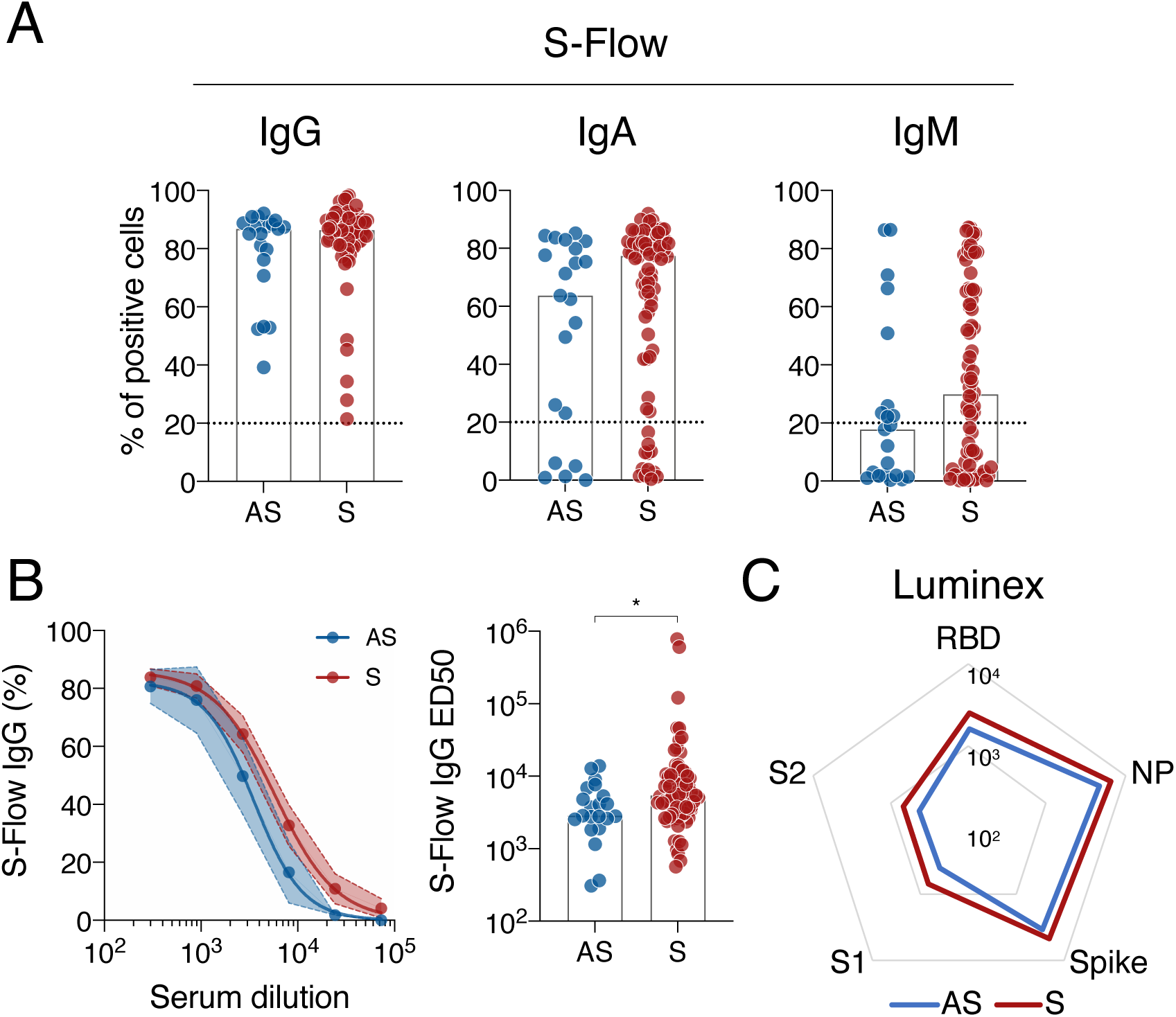
Antibody response to SARS-CoV-2 in sera of asymptomatic and mildly symptomatic COVID-19 individuals. **A**. IgG (left), IgA (middle) and IgM (right) levels were quantified in asymptomatic (AS; blue; n=22) and mildly symptomatic (S; red; n=76) individuals using the flow-cytometry-based S-Flow assay. The percentage of positive cells is represented. **B**. Dose-response analysis of anti-Spike IgG response in AS and S patients measured with the S-Flow assay. Left panel: the mean binding percentage at each serum dilution is represented with the 95% confidence interval. Right panel: the antibody titers (Effective Dose 50%; ED50) of each individual are represented. **C**. Luminex analysis of the antibody response against receptor binding domain (RBD), S1 and S2 subdomains of the Spike, full Spike and nucleoprotein (NP) for AS (blue line) and S (red line) sera. The median of MFI for AS and S sera are represented. In A, the dotted line represents the positivity threshold measured with pre-pandemic sera. In A and C, a Mann-Whitney test was performed (*p<0.05) and the bar indicates the median.

To further document the function of anti-SARS-CoV-2 antibodies, we measured the capacity of sera to neutralize lentiviral Spike pseudotypes in 293T-ACE2 cells (pseudo-neutralization) (Grzelak et al., 2020) and SARS-CoV-2 virus in S-Fuse cells (neutralization) (Buchrieser et al., 2020). We also quantified complement deposition and ADCC activities using the two Raji-Spike-based assays. Sera were interrogated with serial dilutions to calculate exact titers for these four functions. Most of the sera from AS and S individuals were active in these assays **(Figure 4)**. Sera from AS display titers slightly below those from S **(Figure 4A)**. The differences in ID50 were however not significant **(Figure 4B, C)**. We also compared the maximal level of the response **(Figure 4C)**. The level of antibody-mediated complement activation was significantly higher in S than in AS (p=0.036; Mann-Whitney test) **(Figure 4C)**.

**Figure 4.**
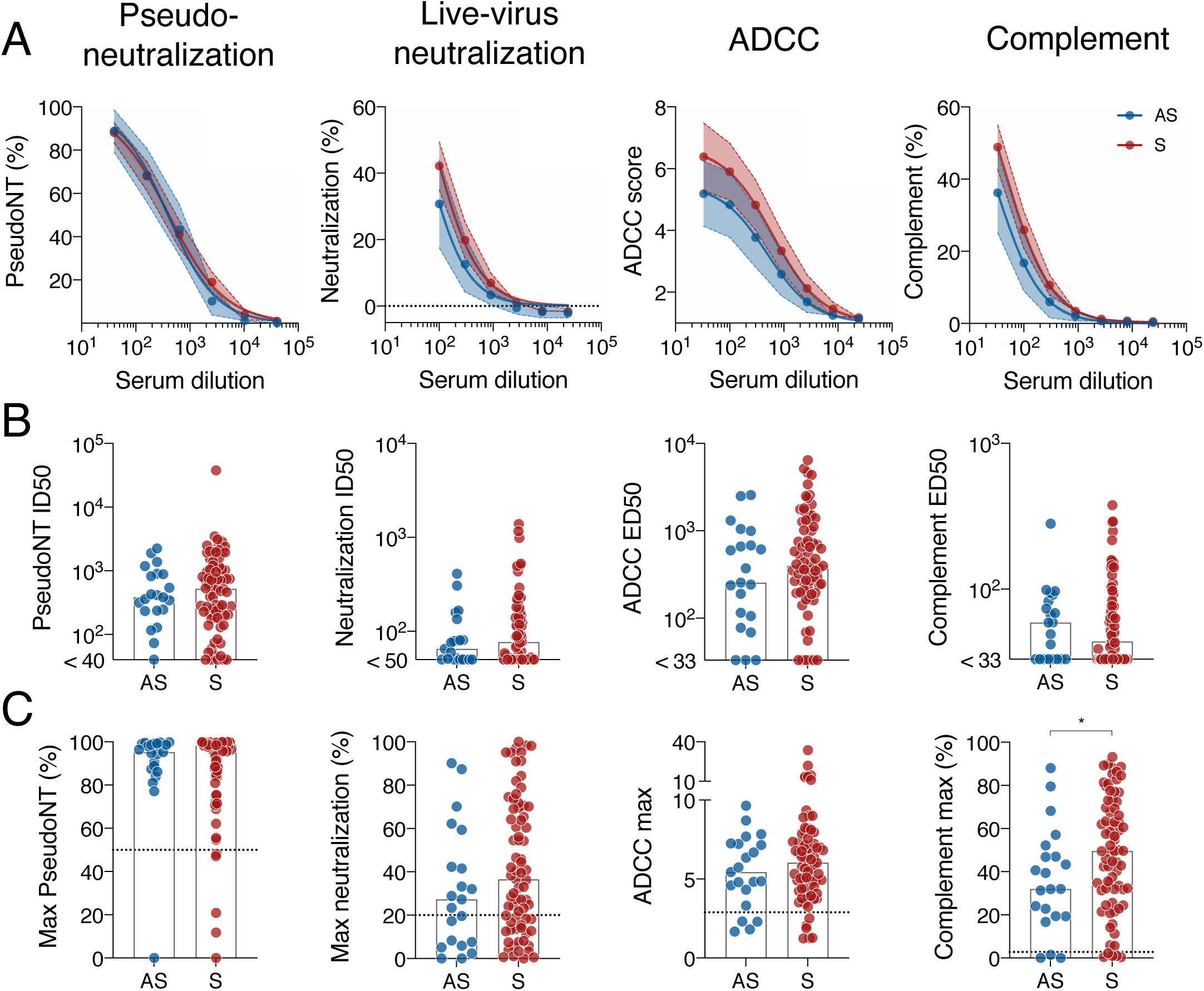
Functional characterization of COVID-19 individuals’ sera. **A**. Asymptomatic (AS; blue) and mildly symptomatic (S; red) sera were tested with serial dilutions for their ability to neutralize Spike pseudoparticles, neutralize SARS-CoV-2, trigger ADCC in the Jurkat-CD16-NFAT-rLuc/Raji-Spike system or induce CDC of Raji-Spike cells. The mean activity at each serum dilution with the 95% confidence interval is depicted for each assay. **B**. The Inhibitory Dilution 50% (ID50) of pseudoneutralization and neutralization, and the Effective Dilution 50% (ED50) of ADCC and CDC induction are depicted. Asymptomatic (AS; blue) and mildly symptomatic (S; red) groups are compared. **C**. The maximum activity of each assay is compared for all asymptomatic (AS; blue) and symptomatic (S; red) individuals. *p<0.05 (Mann-Whitney test). The dotted line on the maximum activity graphs represents the positivity threshold measured with pre-pandemic sera. The bar indicates the median. In B and C, each dot is an individual.

Altogether, our data show that sera of SARS-CoV-2-infected individuals harbor polyfunctional antibodies. Asymptomatic individuals have a decreased capacity to activate the complement and lower IgG titers.

### Correlations between antibody antiviral activities

We then sought to perform an unsupervised analysis of antibody features measured in symptomatic and asymptomatic individuals. We first created correlation matrices of antibody characteristics in each group **(Figure 5A)**. The response to antigens from other viruses and hCoV did not correlate to SARS-CoV-2-related antibody properties. The features corresponding to SARS-CoV-2 appeared positively correlated in both groups. Anti-influenza A IgG measured by Luminex negatively correlated with some SARS-CoV-2 features in AS, but correlation coefficients remained low. ADCC ED50 and maximum activity more strongly correlated to other features in AS compared to S. The IgM-associated features (IgM MFI and IgM %) appeared more strongly correlated to other features in the S group **(Figure 5A)**. Altogether, this analysis reveals slight differences in ADCC and IgM responses between AS and S groups, with an overall high level of coordination in each group. Consistently, Principal Component Analysis (PCA) failed to separate individuals according to their symptoms, showing that no combination of antibody features allowed a strict segregation of the two groups **(Figure 5B)**. This observation was confirmed by unsupervised hierarchical clustering **(Supplementary Figure 4)**.

**Figure 5.**
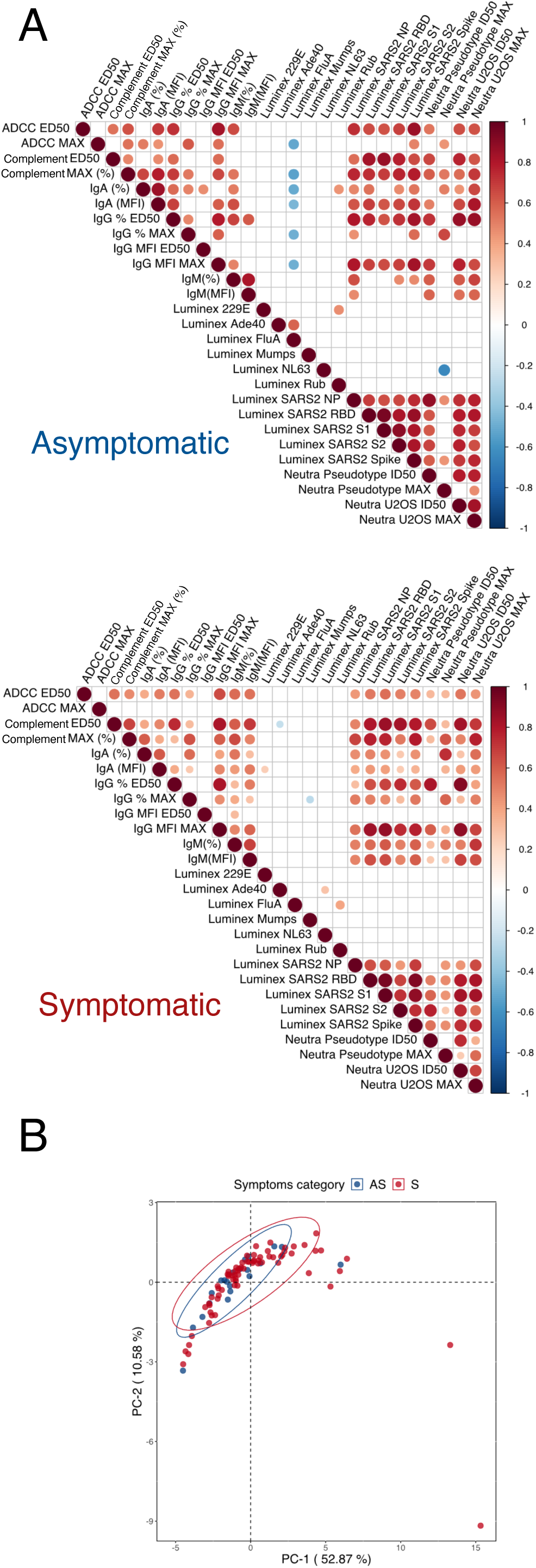
Similarity of antibody response in asymptomatic and symptomatic individuals. **A**. Pearson correlation matrix of all features assessed in both asymptomatic (top) and symptomatic (bottom) individuals. Only statistically significant correlations (p<0.05) are depicted. Antibody features are alphabetically clustered. Positive correlations are indicated in red and negative correlations in blue. The size of the dot is proportional to the p-value. **B**. Principal component analysis of asymptomatic (blue) and symptomatic (red) patients. Each point represents a single patient. The ellipses indicate the Student’s t-distribution with 95% probability.

Overall, this unsupervised analysis shows that the antibody response to SARS-CoV-2 is coordinated in AS and S groups, with differences in ADCC and IgM responses.

### Antibody response to SARS-CoV-2 in other groups of asymptomatic, symptomatic and hospitalized individuals

To further characterize the functionality of antibodies in COVID-19 patients, we analyzed a second cohort established in primary schools of Crépy-en-Valois (n=1340; (Fontanet et al., 2020b) and **Table 2)**. This cohort included a large proportion of children (6-11 years old). We selected all asymptomatic individuals (AS; n=31) and formed a group of sex- and age-matched mildly symptomatic individuals (S; n=43 among the 107 of the cohort) **(Supplementary Figure 5A)**. We also included COVID-19 hospitalized individuals from the FrenchCOVID cohort (H; n=21). As expected, when compared to AS and S, hospitalized patients were older, more often male and sampled sooner after onset of symptoms **(Table 2 and supplementary Figure 5B)**.

**Table 2.**
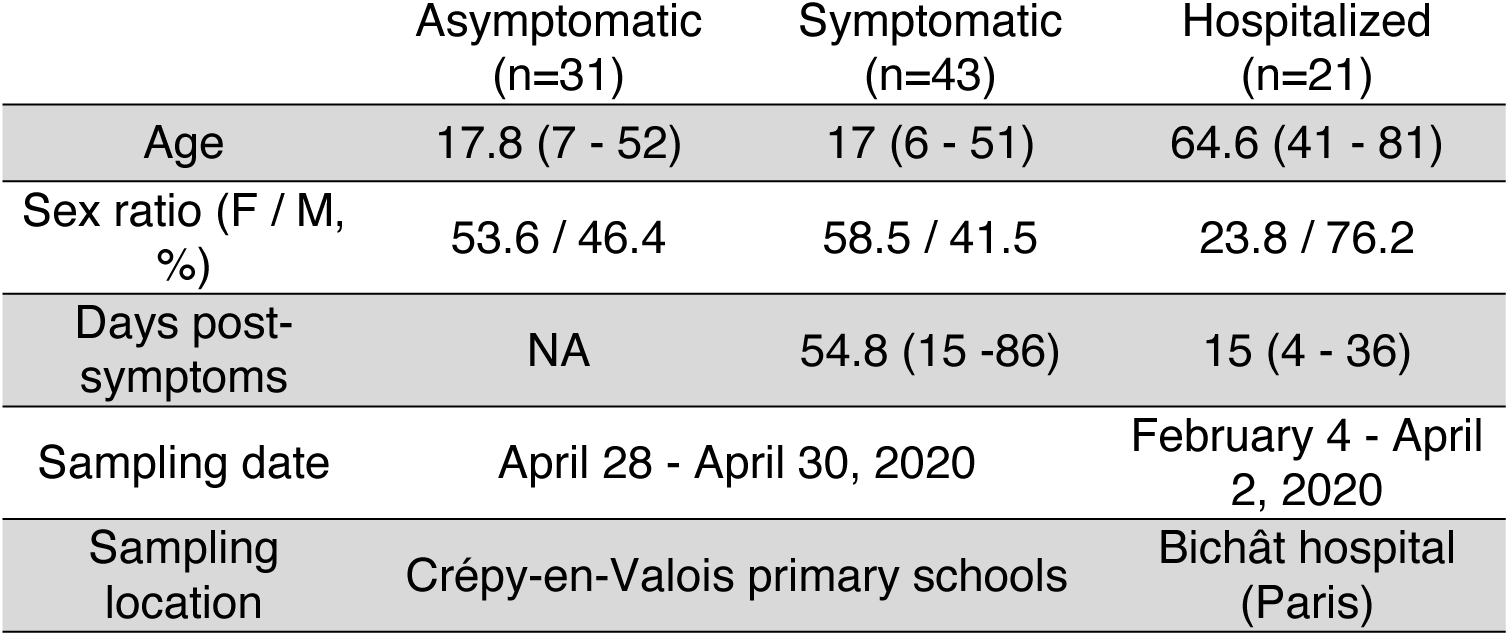
Characteristics of the second group of asymptomatic, symptomatic and hospitalized individuals.

We assessed the anti-Spike antibody profile by S-Flow in the 105 individuals forming these three groups (AS, S and H) **(Figure 6A)**. Frequencies of IgG, IgA and IgM were significantly higher in hospitalized patients than in AS (p=0.0101, p<0.0001 and p<0.0001, Kruskal-Wallis test). When compared to S individuals, hospitalized patients harbored significantly more IgA and IgM (p=0.0002 and p<0.0001, Kruskal-Wallis test). **(Figure 6A**). Measuring the MFI of binding of IgG-, IgA- and IgM-positive individuals showed higher IgG response in hospitalized patients compared to AS and higher IgA response compared to S **(Supplementary Figure 6A)**. We also performed a restricted analysis to compare only AS and S individuals. The frequency of IgA and IgM positive cells, and the MFI of IgG positive cells were significantly higher in S individuals (p=0.0007, p=0014, p=0.018, respectively; Mann-Whitney test) **(Supplementary Figure 6B)**.

**Figure 6.**
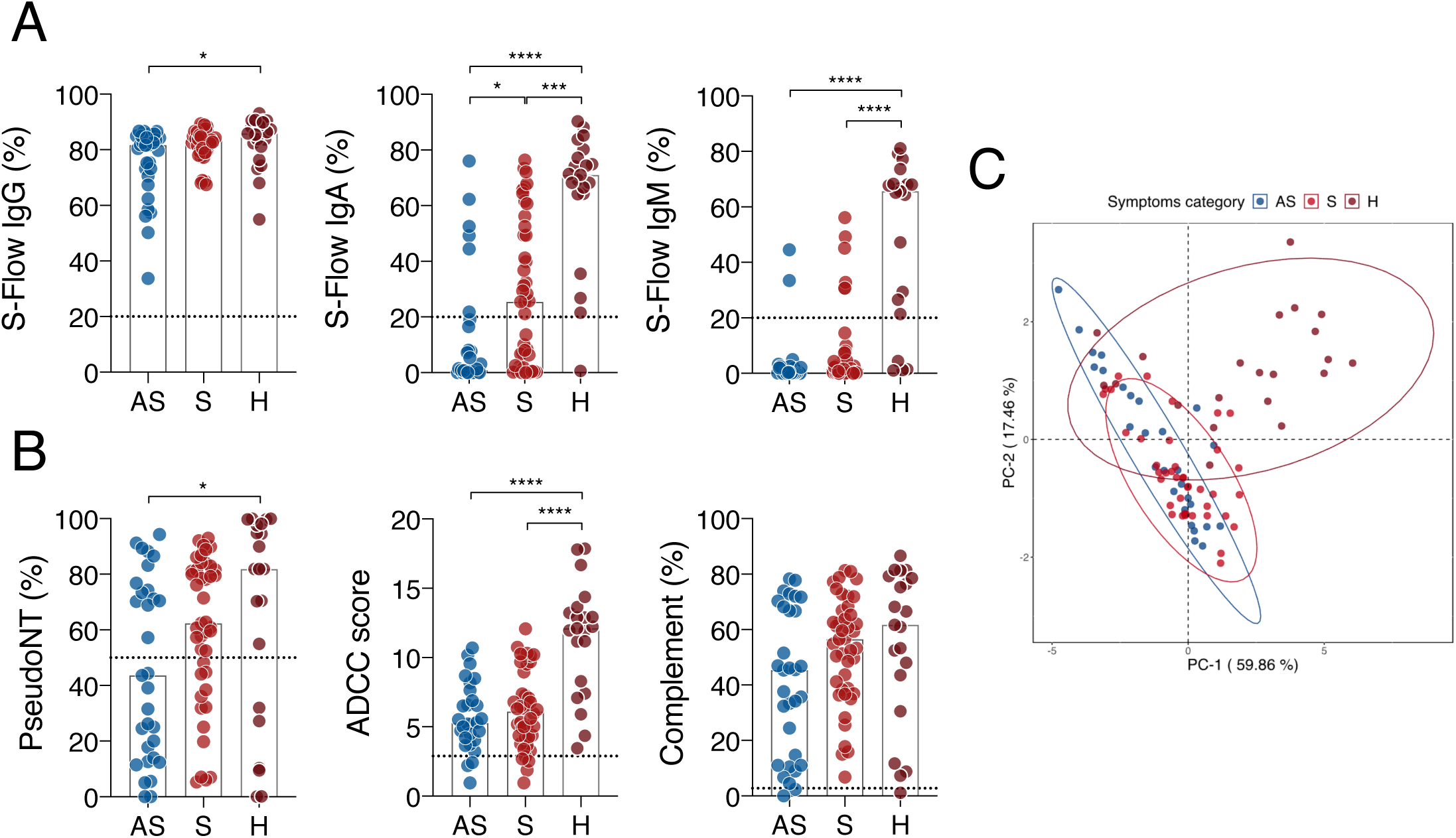
Similarity of antibody response in other groups of asymptomatic, symptomatic and hospitalized individuals. **A**. IgG (left), IgA (middle) and IgM (right) levels were quantified in asymptomatic (AS), mildly symptomatic (S) and hospitalized (H) individuals using the S-Flow assay. The percentage of positive cells is represented. **B**. AS, S and H sera were tested for their ability to neutralize Spike pseudoparticles (left), trigger ADCC in the Jurkat-CD16-NFAT-rLuc/Raji-Spike system (middle) or trigger CDC of Raji-Spike cells (right). **C**. Principal component analysis of asymptomatic (blue), symptomatic (red) and hospitalized (brown) patients. Each point represents a single patient. The ellipses indicate the Student’s t-distribution with 95% probability for each group. In A and B, the dotted line represents the positivity threshold measured with pre-pandemic sera and the bar represents the median. In A and B, a Kruskal-Wallis test was performed (*p<0.05, ***p<0.001, ****p<0.0001).

We then measured pseudo-neutralization, ADCC and complement deposition capacity of the 105 sera. Antibody functions paralleled that of antibody binding, with the highest and lowest activities in hospitalized and asymptomatic individuals, respectively **(Figure 6B)**. Mildly symptomatic individuals scored between hospitalized and asymptomatic individuals **(Figure 6B)**. S resembled AS patients for the level of ADCC, with a lower activity than hospitalized patients (p<0.0001, Kruskal-Wallis test), but were similar to hospitalized individuals for complement deposition **(Figure 6B)**.

To further characterize the anti-Spike response across these 3 groups, we performed unsupervised PCA analysis including the nine antibody features. AS and S clustered together as previously observed in the first groups. In contrast, H individuals appeared divergent, with most individuals clustering apart on the first component (PC-1) **(Figure 6C)**. The contributions of the 9 features to PC-1 ranged from 8% to 14%, indicating that H individuals had an overall higher response, rather than an increase in a limited set of features **(Supplementary Figure 6C)**.

We then mixed the different groups of individuals that we separately analyzed, in order to increase the power of our statistical tests. We thus compared the antibody response of all AS and S individuals (n=52 and n=119, respectively) (Supplementary Figure 7). Frequencies of IgA and IgM were significantly lower in AS than in S (p=0.005, p<0.0001 and p<0.0001, respectively; Mann-Whitney test). Moreover, the MFI of IgG binding, as well as neutralization and complement activities, were significantly lower in AS than in S (p=0.008; Mann-Whitney test. ADCC activity was also lower but did not reach significance.

**Figure 7.**
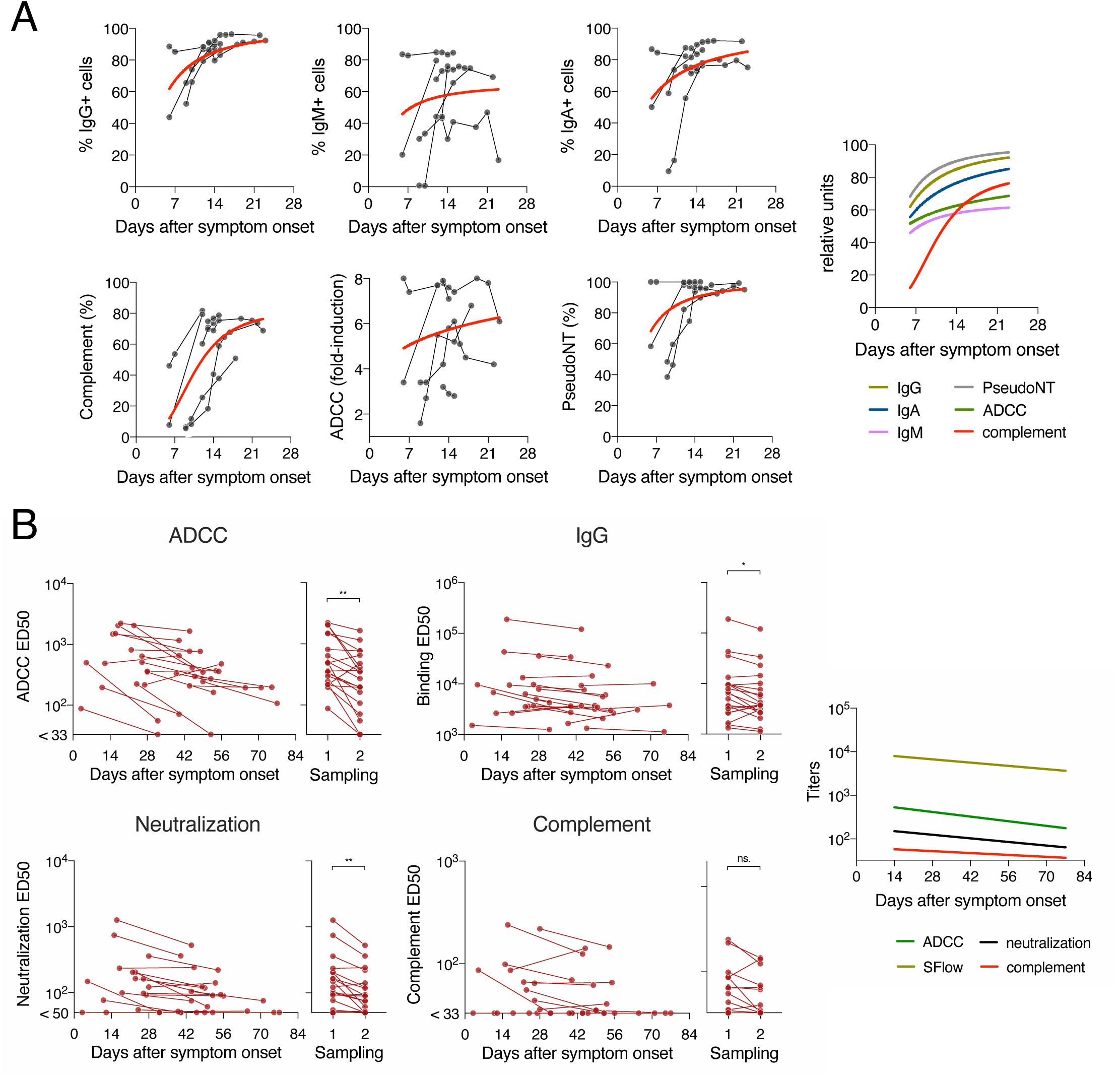
Kinetics of antibody functions in acute hospitalized patients and convalescent mildly symptomatic individuals. **A**. Hospitalized patients (n=7) in the acute phase were sampled at several times post-symptom onset. Their IgG (upper left), IgM (upper middle) and IgA (upper right) levels were assessed longitudinally using the S-Flow assay. CDC (lower left), ADCC (lower middle) and pseudoneutralization (lower right) were also measured. The red curve represents a non-linear fit (4-parameters) of the mean of all donors. The average fit of each function is also depicted (right panel). **B**. Sera from mildly symptomatic patients in the convalescent phase (n=21-22) were sampled twice at a 25-29 days interval. For each serum sample, ADCC (upper left), IgG (upper right), pseudo-neutralization (middle left), real-virus neutralization (middle right) and CDC (lower left) levels were measured with sera serial dilutions. For each assay, the ED50 measured in each assay is represented against the day post-symptom onset on the left graph. On the right graph, data for first and second samplings were pooled in order to perform statistics. *p<0.05, **p<0.01, ***p<0.001, ns. not significant (Wilcoxon test).

The results that asymptomatic individuals harbor a polyfunctional anti-Spike antibody response which is lower than is symptomatic individuals were thus confirmed in a second group of infected persons

### Kinetics of antibody functions after SARS-CoV-2 infection

We then aimed at determining the dynamics of antibody functions after SARS-CoV-2 infection. We identified 7 hospitalized patients and 20 mildly symptomatic COVID-19 cases with longitudinal sampling in our cohorts **(Tables 1 and 2)**. As only symptomatic individuals were invited for a second sampling in our sero-epidemiological studies, no longitudinal samples were available for asymptomatic individuals. Hospitalized patients had the highest temporal resolution and were sampled early after onset of symptoms, with up to 8 samples per individual, from day 6 to 21 post-symptoms onset **(Supplementary Figure 5B)**. We thus documented the induction of the polyfunctional response in these hospitalized patients **(Figure 7A)**. Mildly symptomatic individuals were sampled twice, with a 25-29 days interval. The first sampling was usually performed 21 after onset of symptoms. The mildly symptomatic individuals allowed us to assess the contraction of the immune responses **(Figure 7B)**. We measured IgG, IgA and IgM levels, ADCC, complement deposition, and neutralization or pseudo-neutralization, in the longitudinal samples available.

In hospitalized patients, all functions, except complement deposition, were induced simultaneously between 6 and 21 days after onset of symptoms (**Figure 7A)**. This delay may reflect a lower sensitivity of the assay, a slower appearance of complement-potent antibodies, or a requirement of high antibody titers to trigger this activity.

In mildly symptomatic individuals, we observed a significant decrease in antibody binding titers and their neutralization and ADCC function over time (p=0.011, p=0.001, p=0.001, respectively; Wilcoxon test) **(Figure 7B)**. A decline of complement deposition activity was also visible over time but this was not significant.

## Discussion

We analyzed here different features of the anti-SARS-CoV-2 humoral immune response in infected individuals. We quantified anti-Spike IgG, IgM and IgA as well as antibodies targeting N, S1, S2 and RBD domains, we assessed the amounts of antibodies against two seasonal coronaviruses (229E and NL63) and other viruses (Flu, Mumps, Adenovirus 40 and Rubella). From a functional standpoint, we measured the neutralization activity of the sera using both, lentiviral particles coated with the Spike, and infectious SARS-CoV-2. We also designed different cellular assays to measure the ability of anti-SARS-CoV-2 antibodies to trigger complement deposition and to eliminate infected cells through NK-dependent ADCC. This panel of assays allowed us to establish a profile of the antiviral properties of antibodies in different categories of patients. We focused our analysis on asymptomatic individuals, since their humoral response remains poorly characterized, and compared them to mildly symptomatic and hospitalized patients. We analyzed two groups of asymptomatic and symptomatic individuals, and one group of hospitalized patients, representing a total of 192 individuals.

Overall, we show that asymptomatic individuals mount a polyfunctional anti-SARS-CoV-2 humoral response. The levels of antibodies are lower in asymptomatic individuals, and as a consequence their neutralizing activity and Fc-mediated functions are also reduced. The differences are however modest. Our results were consistent across the two cohorts, despite minor differences such as lower IgA and IgM levels in asymptomatic individuals in the second cohort, when compared to the first one. These differences might be attributed to various sampling times after onset of symptoms and to age variations. Combining AS and S individuals from the two cohorts provided comparable results, albeit differences were more significant, probably as a consequence of increased statistical power. One limitation of our study is the absence of information regarding the date of virus acquisition in asymptomatic individuals. However, both S and AS were sampled at the beginning of the epidemic in Northern France and were likely infected within a short time frame. Our results confirm and extend reports of decreased antibody titers in asymptomatic individuals (Long et al., 2020; Sekine et al., 2020), in which Fc-associated antibody functions were not analyzed. Another limitation is the lack of PCR confirmation for the asymptomatic and mildly symptomatic individuals. Our analysis is therefore restricted to individuals who have seroconverted. Moreover, the different groups were not all age matched. In our first group, the asymptomatic individuals, were as expected, younger than the symptomatic individuals. This was balanced in our second group, in which we selected symptomatic individuals to match the age of the asymptomatic group. It will be of interest to accurately determine the influence of age and other clinical and biological characteristics on the intensity and polyfunctionality of the antibody response.

The levels of antibodies targeting other viruses, including two seasonal coronaviruses were similar in asymptomatic and symptomatic individuals, suggesting that these previous infections did not impact disease severity. One noticeable exception was a higher titer of anti-Flu antibodies in symptomatic persons. The reasons for this remain to be characterized, but a simple explanation may be related to the older age of symptomatic individuals.

We also observed that hospitalized individuals display high levels of anti-SARS-CoV-2 specific antibodies. It has been reported that antibody binding and neutralization are higher in severe and critical cases (Grzelak et al., 2020; Peng et al., 2020; Robbiani et al., 2020; Wang et al., 2020). We further show that complement deposition and ADCC activities are also elevated in hospitalized patients. It has been reported an increase in soluble C5a levels proportional to COVID-19 severity and high levels of C5aR1 expression in blood and pulmonary myeloid cells (Carvelli et al., 2020). The origin of the activation of the C5a-C5aR1 axis in severe COVID-19 remains unknown (Risitano et al., 2020). Whether antibody-mediated complement activation at the surface of infected cells participates to disease severity will deserve further investigation.

It is noteworthy that the neutralizing activity of antibodies correlated with their ability to mediate complement deposition and ADCC, irrespectively of the severity of the disease. A pilot longitudinal analysis performed in some of the hospitalized patients further demonstrated that the acquisition of the different functions similarly increased overtime. The ability to trigger complement deposition was however delayed by one week, compared to ADCC and neutralization. This may be due to a lower sensitivity of our complement test, or to the fact that complement deposition necessitates higher antibody titers than the other functions. By analyzing mildly symptomatic patients at two time points, sampled up to 70 days post symptom onset, we also observed a parallel decline of antibody levels and function. This suggests that neutralization and Fc-mediated activities are performed by the same antibodies, and/or that the half-lives of neutralizing and non-neutralizing antibodies are similar. An analysis of monoclonal antibodies derived from convalescent patients will help determining whether the different functions can be dissociated and may depend on the epitope recognized by individual antibodies.

What are the consequences of complement deposition at the plasma membrane? We show here that sera from COVID-19 patients readily triggered C3 deposition at the surface of infected S-Fuse or A549-ACE2 cells, but C3 deposition did not induce detectable cell death. This may be due to the presence of molecules such as CD59, CD55 and CD46 that counteract complement dependent killing. Raji-Spike cells, which lack CD59 (Dufloo et al., 2019), were indeed readily killed after complement deposition. In contrast, infected S-Fuse cells were rapidly eliminated by NK cells, when anti-SARS-CoV-2 antibodies were added to the culture medium. Our results suggest that NK cells are more potent than complement to kill SARS-CoV-2-infected cells after activation by antibodies. This is in line with previous observations with other viruses. For instance, HIV-1-infected cells may be eliminated by NK cells but not by CDC (Bredow et al., 2016; Bruel et al., 2016; Dufloo et al., 2019). In COVID-19 patients, the main cellular targets of SARS-CoV-2 are ciliated cells from the airways and type II alveolar pneumocytes (Hou et al., 2020). The viral receptor ACE2 is expressed in other tissues than the respiratory tract, and several studies have demonstrated that SARS-CoV-2 has a large cellular tropism (Chu et al., 2020; Hui et al., 2020). Levels of molecules regulating CDC and ADCC vary among cell types (Han et al., 2020; Vivier et al., 2004). Thus, it is likely that these cells display different susceptibilities to ADCC and CDC. Future work will help understanding the sensitivity of natural target cells to anti-viral antibodies, the fate and role of infected cells coated with complement, and the overall contribution of non-neutralizing antibody functions to the immune response to SARS-CoV-2.

Vaccines under development aim at producing neutralizing anti-Spike antibodies (Krammer, 2020). The Fc region is required for optimal efficacy of anti-Spike monoclonal antibodies *in vivo* through mechanisms that may involve those described here (Schäfer et al., 2020). Non-neutralizing antibodies participate to protection offered by experimental vaccines against influenza or HIV (Alter et al., 2020; Dunand et al., 2016). It will be of interest to assess whether SARS-CoV-2 vaccine candidates elicit non-neutralizing antibody functions, and whether these function correlates with vaccine efficacy.

In conclusion, we show here that asymptomatic individuals mount a humoral immune response only slightly decreased when compared to symptomatic SARS-CoV-2 infections. This response includes, in addition to neutralization, the ability to trigger ADCC and complement deposition. Our results warrant further analysis of neutralization and other antibody functions in the evaluation of vaccine candidates and the study of COVID-19 immunopathology.

## Methods

### Cells and viruses

#### Cells

Raji cells (ATCC® CCL‐ 86™) were grown in complete RPMI medium (10% Fetal Calf Serum (FCS), 1% Penicillin/Streptomycin (PS)). 293T cells (ATCC® CRL‐ 3216™) and U2OS cells (ATCC® HTB‐ 96™) were grown in complete DMEM medium (10% FCS, 1% PS). A549 cells (ATCC® CCL‐ 185™) were cultured in F-12K Nutrient Mixture Media with 10% FCS and 1% PS. 293T, U2OS and A549 cells stably expressing ACE2, and U2OS-ACE2 cells stably expressing the GFPsplit system (GFP1-10 and GFP11; S-Fuse cells) were previously described (Buchrieser et al., 2020). Blasticidin (10 μg/mL) and puromycin (1 μg/mL) were used to select for ACE2 and GFPsplit transgenes expression, respectively. 293T and Raji cells stably expressing the SARS-CoV-2 Spike protein (GenBank: QHD43416.1) were generated by lentiviral transduction and selection with puromycin (1 μg/mL). Absence of mycosplasma contamination was confirmed in all cell lines with the Mycoalert™ Mycoplasma Detection Kit (Lonza).

#### Viruses

The SARS-CoV-2 strain BetaCoV/France/IDF0372/2020 was supplied by the National Reference Centre for Respiratory Viruses hosted by Institut Pasteur (Paris, France). The human sample from which the strain was isolated has been provided by Dr. X. Lescure and Pr. Y. Yazdanpanah from the Bichât Hospital, Paris, France. The viral strain was supplied through the European Virus Archive goes Global (Evag) platform (Horizon 2020 research and innovation grant n°653316). Titration of viral stocks was performed on Vero E6, with a limiting dilution technique allowing a calculation of PFU (plaque-forming unit)/ml.

### Human samples

#### Pre-pandemic individuals’ sera

Pre-pandemic sera were randomly sampled from 200 anonymized healthy blood donors recruited in March 2017 at the Val d’Oise sites of EFS (the French blood agency). The ICAReB platform (BRIF code n°BB-0033-00062) of Institut Pasteur collects and manages bioresources following ISO (International Organization for Standardization) 9001 and NF S 96-900 quality standards.

#### COVID-19 asymptomatic and mildly symptomatic convalescent sera

Following the first documented local transmission of SARS-CoV-2 in France, outbreak investigation and contact tracing identified two cases in the high school of Crépy-en-Valois (France) on 2 February 2020. We conducted two retrospective seroepidemiological studies in the city:

i. First, a retrospective closed cohort study in the high school (Fontanet et al., 2020a). Between 30 March and 4 April, all pupils, as well as teachers and non-teaching staff (administrative, cleaners, catering) from the high school were invited to participate in the investigation (n=1200). Participants completed a questionnaire which covered sociodemographic information and a 5 mL blood sample was taken (n=661). Some of the participants (n=203) had a previous blood sampling as part of an initial investigation of the cluster on March 3-4, 2020.
ii. Second, an investigation across primary schools (Fontanet et al., 2020b). We invited all pupils, teachers and non-teaching staff (administrative, cleaners, catering) from each of the six primary schools who were registered at the school from the beginning of the epidemic (estimated around 13 January 2020) up to the time of the investigation on April 28-30, 2020. Participants (with the help of their parents in the case of pupils) completed a questionnaire that covered sociodemographic information and a 5 mL blood sample was taken (n=1340).

These studies are registered with Clinicaltrials.gov (ID: NCT04325646) and received ethical approval by the Comité de Protection des Personnes Ile-de-France (CPP-IDF) III. Informed consent was obtained from all study participants. The ICAReB platform (BRIF code n°BB-0033-00062) of Institut Pasteur collected and managed bioresources following ISO (International Organization for Standardization) 9001 and NF S 96-900 quality standards.

#### Sera from hospitalized COVID-19 patients

Serum samples from hospitalized COVID-19 cases were obtained from Hôpital Bichat–Claude-Bernard as part of the French COVID-19 cohort. Each participant provided written consent to participate in the study, which was approved by the regional investigational review board (CPP-IDF VII, Paris, France) (ID RCB: 2020-A00256-33) and performed according to European guidelines and the Declaration of Helsinki.

#### Healthy blood donors for NK cells isolation

Peripheral blood mononuclear cells (PBMCs) were isolated from peripheral blood of healthy human donors from the Etablissement Français du Sang (EFS), in accordance with local ethical guidelines. NK cells were enriched by magnetic negative selection (Miltenyi) and cultured overnight in complete RPMI medium before use.

### Serological analysis of antibodies (S-Flow and Luminex)

#### S-Flow

The S-Flow assay was performed as previously described (Grzelak et al., 2020). Briefly, 293T-S were incubated at 4°C for 30 min with sera (1:300 dilution, unless otherwise specified) in PBS containing 0.5% BSA and 2 mM EDTA, washed with PBS, and stained using either anti-IgG Alexa Fluor 647 (dilution 1:600; Thermo Fisher Scientific), or anti-IgM Alexa Fluor 488 (dilution 1:600; Thermo Fisher Scientific), or anti-IgA Alexa Fluor 647 (dilution 1:800; Jackson ImmunoResearch). Cells were washed with PBS and fixed for 10 min using 4% paraformaldehyde (PFA). Data were acquired on an Attune NxT instrument (Life Technologies). Specific binding was calculated with the following formula: 100 × (% binding on 293T-Spike − % binding on control cells)/(100 − % binding on control cells).

#### Luminex

A multiplex Luminex^®^ MAGPIX^®^ assay was developed to measure IgG antibody responses to SARS-CoV-2 antigens (trimeric Spike, S1, S2 RBD, Nucleoprotein), Nucleoprotein from two seasonal coronaviruses (NL63 and 229E), and antigens from other viruses (Influenza A H1N1, adenovirus type 40, mumps, rubella)(Rosado et al., 2020).

### Antibody-dependent cellular cytotoxicity (ADCC) assays

#### ADCC assay on infected cells

4×10^3^ U2OS-ACE2-GFP-1-10 and 4×10^3^ U2OS-ACE2-GFP-11 cells were plated in a μClear 96-well plate (Greiner Bio-One). The next day, cells were infected at a multiplicity of infection (MOI) of 0.1 for 18h. NK cells isolated from PBMCs of a healthy donor (ratio 1:1) and sera from pre-pandemic or COVID-19 individuals (dilution 1:100) were added. After 4 hours, cells were fixed with 2% PFA, washed, stained with Hoechst (dilution 1:1,000, Invitrogen) and acquired with an Opera Phenix high content confocal microscope (PerkinElmer). The GFP area was quantified with the Harmony software (PerkinElmer). ADCC was measured using the formula: 100 x (GFP area in “no serum” – GFP area in “tested serum”)/(GFP area in “no serum”).

#### CD16 activation reporter assay

ADCC was quantified using the ADCC Reporter Bioassay (Promega) according to manufacturer’s instructions. Briefly, 5×10^4^ Raji-Spike cells were co-cultured with 5×10^4^ Jurkat-CD16-NFAT-rLuc cells in presence or absence of pre-pandemic or COVID-19 sera at the indicated dilution. Luciferase was measured after 18 hours of incubation using an EnSpire plate reader (PerkinElmer). ADCC was measured as the fold induction of Luciferase activity compared to the “no serum” condition.

### Complement activation assays

#### Complement activation assay on infected cells

1.5×10^4^ A549-ACE2 cells were plated in 96-well plates. After overnight incubation, cells were infected at a MOI of 1 for 24 hours. Then, pre-pandemic or COVID-19 serum was added as a source of antibodies (dilution 1:100) and normal (NHS) or heat-inactivated (HIHS) serum was added as a source of complement (dilution 1:2). After 4 hours, cells were detached using PBS-EDTA and incubated 30 min at 4°C with an APC‐conjugated anti‐ C3b/iC3b antibody (clone 6C9, Tebu‐bio, dilution 1:50). Cells were washed with PBS and fixed with 4% PFA. For Spike staining, cells were incubated 30 min with biotinylated anti-Spike antibody (10 μg/mL in PBS/BSA 0.5%/Saponin 0.05%), washed, and incubated 30 min with Streptavidin R-PE (dilution 1:100 in PBS/BSA 0.5%/Saponin 0.05%, Invitrogen). Anti-Spike antibody was a kind gift of Hugo Mouquet (Institut Pasteur, Paris). Data were acquired on an Attune NxT instrument (Life Technologies). For each serum, complement-dependent cytotoxicity (CDC) of infected cells was calculated using the following formula: 100 × (% of infected cells with HIHS − % of infected cells with NHS)/(% of infected cells with HIHS).

#### Complement-dependent cytotoxicity (CDC) on Raji-Spike cells

CDC of Raji cells was measured as previously described (Dufloo et al., 2019). Briefly Raji-Spike cells (5×10^4^) were cultivated in the presence of 50% normal (NHS) or heat‐inactivated (HIHS) human serum and with or without pre-pandemic or COVID-19 sera (diluted 1:33 unless otherwise stated). After 24h, cells were washed with PBS and the live/dead fixable aqua dead cell marker (1:1,000 in PBS; Life Technologies) was added for 30 min at 4°C before fixation. Data were acquired on an Attune NxT instrument (Life Technologies). CDC was calculated using the following formula: 100 × (% of dead cells with serum − % of dead cells without serum)/(100 − % of dead cells without serum).

### Neutralization assays

#### Pseudo-neutralization assay

2×10^4^ 293T-ACE2 cells were plated in 96-well plates. Single cycle lentiviral Spike pseudotypes encoding for a luciferase reporter gene were preincubated 30 minutes at room temperature with the serum to be tested at the indicated dilution and added to the cells. The luciferase signal was measured after 48 h. The percentage of neutralization was calculated with the following formula, setting the “no serum” condition at 0% and the “no-pseudotype” condition at 100%: 100 x (1 – (value with serum – value with “no-pseudotype”)/(value with “no serum” – value with “no-pseudotype”)).

#### SARS-CoV-2 neutralization assay (S-Fuse)

4×10^3^ U2OS-ACE0-GFP-1-10 and 4×10^3^ U2OS-ACE0-GFP-11 cells were plated overnight in a μClear 96-well plate (Greiner Bio-One). SARS-CoV-2 was incubated with sera at the indicated dilutions for 30 minutes at room temperature and added on S-fuse cells (MOI 0.1). 18 hours later, cells were fixed with 2% PFA, washed, stained with Hoechst (dilution 1:1,000, Invitrogen) and acquired with an Opera Phenix high content confocal microscope (PerkinElmer). For each well, the GFP area and the number of nuclei were quantified using the Harmony software (PerkinElmer). The percentage of neutralization was calculated using the nuclei count or the GFP area using the following formula, setting the “no serum” condition at 0% and the non-infected condition at 100%: 100 x (1 – (value with serum – value in “non-infected”)/(value in “no serum” – value in “non-infected”)).

### Live-imaging

1×10^4^ U2OS-ACE0-GFP-1-10 and 1×10^4^ U2OS-ACE0-GFP-11 cells were plated overnight in each compartment of a μ-Dish^35 mm^ Quad (Ibidi). The next day, cells were infected at a MOI of 0.1. 18 hours later, NK cells were added at 1:1 ratio as well as serum from a pre-pandemic or a COVID-19 individual (dilution 1:100). Conditions without NK and Serum (“No serum No NK”) and with NK cells but without serum (“no serum”) were included as controls. Propidium iodide (PI) (10 μg/ml, Life Technologies) was added to monitor cell death. Transmission and fluorescence images were acquired at a 20X magnification every 4 minutes for 4 hours on a BioStation IM-Q (Nikon). At least 5 fields were recorded in each condition. Images were analyzed using the FIJI software.

### Statistical analysis

Calculations, figures and statistics were performed using Excel 365 (Microsoft), Prism 8 (GraphPad Software) or RStudio Desktop 1.3.1093 (R Studio, PBC). For R analysis we used the following packages: corrplot (https://github.com/taiyun/corrplot), pheatmap (https://CRAN.R-project.org/package=pheatmap), factoextra and FactoMineR (https://CRAN.R-project.org/package=factoextra) and readxl (https://CRAN.R-project.org/package=readxl).

## Data Availability

All data are available.

## Acknowledgments

We thank the individuals who donated their blood. We thank Michael Maaran Rajah for critical reading of the manuscript; J. Toubiana and S. Brisse (Institut Pasteur) for providing pre-pandemic serum samples; R. Lemahieu for organizing the epidemiological survey; and the ICAReB team for management and distribution of serum samples. We thank F. Mentre, S. Tubiana, the French COVID-19 cohort, and REACTing for cohort management. We thank Sandrine Ferandes Pellerin and Nathalie Jolly for their coordination of the CORSER studies.

## Author contribution

Conceptualization and Methodology: JD, LG, OS and TB

Cohort management and sample collection: AW, MNU, YL and BH

Acquisition or analysis of data: JD, LG, IS, YM, LT, FA, SP, CP, JB, RR, HM, PC, MW, AF, OS and TB

Data assembly and manuscript writing: JD, LG, OS and TB

Funding acquisition: HM, PC, MW, YL, BH, AF, OS and TB Supervision: OS and TB

## Tables legends

Table 1. Characteristics of patients enrolled in Crépy-en-Valois high school.

Table 2. Characteristics of patients enrolled in Crépy-en-Valois primary schools and at hospitals.

## Supplementary Figures legends

**Figure S1 (related to Figure 1)**.

**A**. Percentage of C3+ cells among infected (Spike+) for each pre-pandemic (left) and COVID-19 (right) serum in presence of normal human serum (NHS) as a source of complement. Each dot represents an independent experiment.

**B**. Percentage of C3+ cells among infected (Spike+) for each pre-pandemic (left) and COVID-19 (right) serum in presence of heat-inactivated human serum (HIHS) as a control. Each dot represents an independent experiment.

**C**. Complement-dependent cytotoxicity (CDC) of infected A549-ACE2 cells was measured for each pre-pandemic (left) and COVID-19 (right) serum as the relative disappearance of Spike+ cells in the NHS condition compared to the HIHS condition. Each dot represents an independent experiment.

**Figure S2 (related to Figure 2)**.

The percentage of ADCC of infected (Spike+) cells was measured for each pre-pandemic (left) and COVID-19 serum (right). Each dot represents a different donor of NK cells (n=2-6).

**Figure S3 (related to Figure 3)**.

**A**. IgG (left), IgA (middle) and IgM (right) levels were quantified in asymptomatic (AS; blue) and mildly symptomatic (S; red) individuals using the flow-cytometry-based S-Flow assay. The median fluorescence intensity (MFI) of staining in S-Flow+ individuals is represented.

**B-D**. Asymptomatic (n=22; AS) and symptomatic (n=76; S) sera were analyzed by Luminex to measure the antibody response against SARS-CoV-2 viral antigens **(B)**, seasonal coronaviruses 229E and NL63 **(C)** and control antigens **(D)**. The median fluorescence intensities (MFI) are represented. The bar represents the median. **p<0.01 (Mann-Whitney test).

**Figure S4 (related to Figure 5)**.

Hierarchical clustering of asymptomatic and symptomatic patients based on serological features. Patients and antibody features were clustered using k-means clustering. Each column represents an individual patient. Each feature (line) was normalized with minimum value in purple and maximum value in orange. Patients were tagged as asymptomatic (blue, 21 individuals) or symptomatic (red, 70 individuals).

**Figure S5 (related to Figure 6 and Table 1 and 2)**.

**A**. The duration post-symptom onset, the age and the sex ratio were compared between the first cohort (AS and S), the second cohort (AS and S) and the hospitalized patients (H). A Kruskal-Wallis test was performed. ***p<0.001, ****p<0.0001.

**B**. The sex ration and the age was compared between asymptomatic and symptomatic patients from the first and second cohorts. A Mann-Whitney test was performed. ns. not significant, **p<0.01.

**Figure S6 (related to Figure 6)**.

**A**. IgG (left), IgA (middle) and IgM (right) levels were quantified in asymptomatic (AS; blue), mildly symptomatic (S; red) and hospitalized (H; brown) individuals using the flow-cytometry-based S-Flow assay. The median fluorescence intensity (MFI) of staining in S-Flow+ individuals is represented. The bar represents the median. *p<0.05, **p<0.01 (Kruskal-Wallis test).

**B**. IgG, IgA and IgM levels in asymptomatic and symptomatic patients were compared using a Mann-Whitney test. Comparisons include the percentages of positive cells in all individuals (grey) and the MFI of staining in S-Flow+ individuals only (blue).

**C**. Contribution percentages to first (left) and second (right) dimensions of the 9 antibody features included in the principal component analysis performed on asymptomatic, symptomatic and hospitalized patients.

**Figure S7 (related to Figure 6)**.

**A**. IgG (left), IgA (middle) and IgM (right) levels were quantified in asymptomatic (AS; blue) and mildly symptomatic (S; red) individuals from the two cohorts using the flow-cytometry-based S-Flow assay. The percentage of positive cells is represented.

**B**. IgG (left), IgA (middle) and IgM (right) levels were quantified in asymptomatic (AS; blue) and mildly symptomatic (S; red) individuals from the two cohorts using the flow-cytometry-based S-Flow assay. The median fluorescence intensity (MFI) of staining in S-Flow+ individuals is represented.

**C**. AS and S sera from the two cohorts were tested for their ability to neutralize Spike pseudoparticles (left), trigger ADCC in the Jurkat-CD16-NFAT-rLuc/Raji-Spike system (middle) or trigger CDC of Raji-Spike cells (right).

The bar indicates the median. In A, the dotted line represents the threshold of positivity measured with pre-pandemic sera. Mann-Whitney tests were performed (*p<0.05; **p<0.01; ***p<0.001; ****p<0.0001).

## References

Alter, G., Yu, W.-H., Chandrashekar, A., Borducchi, E.N., Ghneim, K., Sharma, A., Nedellec, R., McKenney, K.R., Linde, C., Broge, T., et al. (2020). Passive Transfer of Vaccine-Elicited Antibodies Protects against SIV in Rhesus Macaques. Cell 183, 185-196.e14.

Atyeo, C., Fischinger, S., Zohar, T., Slein, M.D., Burke, J., Loos, C., McCulloch, D.J., Newman, K.L., Wolf, C., Yu, J., et al. (2020). Distinct early serological signatures track with SARS-CoV-2 survival. Immunity.

Barnes, C.O., West, A.P., Huey-Tubman, K.E., Hoffmann, M.A.G., Sharaf, N.G., Hoffman, P.R., Koranda, N., Gristick, H.B., Gaebler, C., Muecksch, F., et al. (2020). Structures of human antibodies bound to SARS-CoV-2 spike reveal common epitopes and recurrent features of antibodies. Cell 182, 828-842.e16.

Bastard, P., Rosen, L.B., Zhang, Q., Michailidis, E., Hoffmann, H.-H., Zhang, Y., Dorgham, K., Philippot, Q., Rosain, J., Béziat, V., et al. (2020). Autoantibodies against type I IFNs in patients with life-threatening COVID-19. Science 370, eabd4585.

Blanco-Melo, D., Nilsson-Payant, B.E., Liu, W.-C., Uhl, S., Hoagland, D., Møller, R., Jordan, T.X., Oishi, K., Panis, M., Sachs, D., et al. (2020). Imbalanced Host Response to SARS-CoV-2 Drives Development of COVID-19. Cell 181, 1036-1045.e9.

Bournazos, S., and Ravetch, J.V. (2017). Fcγ Receptor Function and the Design of Vaccination Strategies. Immunity 47, 224–233.

Bournazos, S., Klein, F., Pietzsch, J., Seaman, M.S., Nussenzweig, M.C., and Ravetch, J.V. (2014). Broadly neutralizing anti-HIV-1 antibodies require Fc effector functions for in vivo activity. Cell 158, 1243 1253.

Bredow, B. von, Arias, J.F., Heyer, L.N., Moldt, B., Le, K., Robinson, J.E., Zolla-Pazner, S., Burton, D.R., and Evans, D.T. (2016). Comparison of Antibody-Dependent Cell-Mediated Cytotoxicity and Virus Neutralization by HIV-1 Env-Specific Monoclonal Antibodies. J Virol 90, 6127–6139.

Bruel, T., Guivel-Benhassine, F., Amraoui, S., Malbec, M., Richard, L., Bourdic, K., Donahue, D.A., Lorin, V., Casartelli, N., Noël, N., et al. (2016). Elimination of HIV-1-infected cells by broadly neutralizing antibodies. Nat Commun 7, 10844.

Buchrieser, J., Dufloo, J., Hubert, M., Monel, B., Planas, D., Rajah, M.M., Planchais, C., Porrot, F., Guivel‐Benhassine, F., Werf S.V. der, et al. (2020). Syncytia formation by SARS‐ CoV‐ 2 infected cells. Embo J e2020106267.

Carvelli, J., Demaria, O., Vély, F., Batista, L., Benmansour, N.C., Fares, J., Carpentier, S., Thibult, M.-L., Morel, A., Remark, R., et al. (2020). Association of COVID-19 inflammation with activation of the C5a–C5aR1 axis. Nature 1–9.

Chu, H., Chan, J.F.-W., Yuen, T.T.-T., Shuai, H., Yuan, S., Wang, Y., Hu, B., Yip, C.C.-Y., Tsang, J.O.-L., Huang, X., et al. (2020). Comparative tropism, replication kinetics, and cell damage profiling of SARS-CoV-2 and SARS-CoV with implications for clinical manifestations, transmissibility, and laboratory studies of COVID-19: an observational study. Lancet Microbe 1, e14–e23.

Cummings, M.J., Baldwin, M.R., Abrams, D., Jacobson, S.D., Meyer, B.J., Balough, E.M., Aaron, J.G., Claassen, J., Rabbani, L.E., Hastie, J., et al. (2020). Epidemiology, clinical course, and outcomes of critically ill adults with COVID-19 in New York City: a prospective cohort study. Lancet 395, 1763–1770.

Dai, H.-S., Griffin, N., Bolyard, C., Mao, H.C., Zhang, J., Cripe, T.P., Suenaga, T., Arase, H., Nakano, I., Chiocca, E.A., et al. (2017). The Fc Domain of Immunoglobulin Is Sufficient to Bridge NK Cells with Virally Infected Cells. Immunity 47, 159-170.e10.

DiLillo, D.J., Palese, P., Wilson, P.C., and Ravetch, J.V. (2016). Broadly neutralizing anti-influenza antibodies require Fc receptor engagement for in vivo protection. J Clin Invest 126, 605–610.

Dufloo, J., Guivel‐Benhassine, F., Buchrieser, J., Lorin, V., Grzelak, L., Dupouy, E., Mestrallet, G., Bourdic, K., Lambotte, O., Mouquet, H., et al. (2019). Anti‐ HIV ‐ 1 antibodies trigger non‐lytic complement deposition on infected cells. Embo Rep 21, e49351.

Dunand, C.J.H., Leon, P.E., Huang, M., Choi, A., Chromikova, V., Ho, I.Y., Tan, G.S., Cruz, J., Hirsh, A., Zheng, N.-Y., et al. (2016). Both Neutralizing and Non-Neutralizing Human H7N9 Influenza Vaccine-Induced Monoclonal Antibodies Confer Protection. Cell Host Microbe 19, 800–813.

Fafi-Kremer, S., Bruel, T., Madec, Y., Grant, R., Tondeur, L., Grzelak, L., Staropoli, I., Anna, F., Souque, P., Fernandes-Pellerin, S., et al. (2020). Serologic responses to SARS-CoV-2 infection among hospital staff with mild disease in eastern France. Ebiomedicine 102915.

Flaxman, S., Mishra, S., Gandy, A., Unwin, H.J.T., Mellan, T.A., Coupland, H., Whittaker, C., Zhu, H., Berah, T., Eaton, J.W., et al. (2020). Estimating the effects of non-pharmaceutical interventions on COVID-19 in Europe. Nature 584, 257–261.

Fontanet, A., Tondeur, L., Madec, Y., Grant, R., Besombes, C., Jolly, N., Pellerin, S.F., Ungeheuer, M.-N., Cailleau, I., Kuhmel, L., et al. (2020a). Cluster of COVID-19 in northern France: A retrospective closed cohort study. MedRxiv.

Fontanet, A., Grant, R., Tondeur, L., Madec, Y., Grzelak, L., Cailleau, I., Ungeheuer, M.-N., Renaudat, C., Pellerin, S.F., Kuhmel, L., et al. (2020b). SARS-CoV-2 infection in primary schools in northern France: A retrospective cohort study in an area of high transmission. MedRxiv.

Giamarellos-Bourboulis, E.J., Netea, M.G., Rovina, N., Akinosoglou, K., Antoniadou, A., Antonakos, N., Damoraki, G., Gkavogianni, T., Adami, M.-E., Katsaounou, P., et al. (2020). Complex Immune Dysregulation in COVID-19 Patients with Severe Respiratory Failure. Cell Host Microbe 27, 992-1000.e3.

Gorbalenya, A.E., Baker, S.C., Baric, R.S., Groot R.J. de, Drosten, C., Gulyaeva, A.A., Haagmans, B.L., Lauber, C., Leontovich, A.M., Neuman, B.W., et al. (2020). The species Severe acute respiratory syndrome-related coronavirus: classifying 2019-nCoV and naming it SARS-CoV-2. Nat Microbiol 5, 536–544.

Grzelak, L., Temmam, S., Planchais, C., Demeret, C., Tondeur, L., Huon, C., Guivel-Benhassine, F., Staropoli, I., Chazal, M., Dufloo, J., et al. (2020). A comparison of four serological assays for detecting anti-SARS-CoV-2 antibodies in human serum samples from different populations. Sci Transl Med eabc3103.

Guan, W.-J., Ni, Z.-Y., Hu, Y., Liang, W.-H., Ou, C.-Q., He, J.-X., Liu, L., Shan, H., Lei, C.-L., Hui, D.S.C., et al. (2020). Clinical Characteristics of Coronavirus Disease 2019 in China. New Engl J Med 382, 1708–1720.

Gunn, B.M., Yu, W.-H., Karim, M.M., Brannan, J.M., Herbert, A.S., Wec, A.Z., Halfmann, P.J., Fusco, M.L., Schendel, S.L., Gangavarapu, K., et al. (2018). A Role for Fc Function in Therapeutic Monoclonal Antibody-Mediated Protection against Ebola Virus. Cell Host Microbe 24, 221-233.e5.

Hadjadj, J., Yatim, N., Barnabei, L., Corneau, A., Boussier, J., Smith, N., Péré, H., Charbit, B., Bondet, V., Chenevier-Gobeaux, C., et al. (2020). Impaired type I interferon activity and inflammatory responses in severe COVID-19 patients. Science 369, 718–724.

Han, X., Zhou, Z., Fei, L., Sun, H., Wang, R., Chen, Y., Chen, H., Wang, J., Tang, H., Ge, W., et al. (2020). Construction of a human cell landscape at single-cell level. Nature 581, 303–309.

Hoffmann, M., Kleine-Weber, H., Schroeder, S., Krüger, N., Herrler, T., Erichsen, S., Schiergens, T.S., Herrler, G., Wu, N.-H., Nitsche, A., et al. (2020). SARS-CoV-2 Cell Entry Depends on ACE2 and TMPRSS2 and Is Blocked by a Clinically Proven Protease Inhibitor. Cell 181, 271-280.e8.

Hou, Y.J., Okuda, K., Edwards, C.E., Martinez, D.R., Asakura, T., Dinnon, K.H., Kato, T., Lee, R.E., Yount, B.L., Mascenik, T.M., et al. (2020). SARS-CoV-2 Reverse Genetics Reveals a Variable Infection Gradient in the Respiratory Tract. Cell 182, 429-446.e14.

Huang, C., Wang, Y., Li, X., Ren, L., Zhao, J., Hu, Y., Zhang, L., Fan, G., Xu, J., Gu, X., et al. (2020). Clinical features of patients infected with 2019 novel coronavirus in Wuhan, China. Lancet 395, 497–506.

Hui, K.P.Y., Cheung, M.-C., Perera, R.A.P.M., Ng, K.-C., Bui, C.H.T., Ho, J.C.W., Ng, M.M.T., Kuok, D.I.T., Shih, K.C., Tsao, S.-W., et al. (2020). Tropism, replication competence, and innate immune responses of the coronavirus SARS-CoV-2 in human respiratory tract and conjunctiva: an analysis in ex-vivo and in-vitro cultures. Lancet Respir Medicine 8, 687–695.

Krammer, F. (2020). SARS-CoV-2 vaccines in development. Nature 1–12.

Lavezzo, E., Franchin, E., Ciavarella, C., Cuomo-Dannenburg, G., Barzon, L., Vecchio, C.D., Rossi, L., Manganelli, R., Loregian, A., Navarin, N., et al. (2020). Suppression of a SARS-CoV-2 outbreak in the Italian municipality of Vo’. Nature 584, 425–429.

Long, Q.-X., Tang, X.-J., Shi, Q.-L., Li, Q., Deng, H.-J., Yuan, J., Hu, J.-L., Xu, W., Zhang, Y., Lv, F.-J., et al. (2020). Clinical and immunological assessment of asymptomatic SARS-CoV-2 infections. Nat Med 26, 1200– 1204.

Lucas, C., Wong, P., Klein, J., Castro, T.B.R., Silva, J., Sundaram, M., Ellingson, M.K., Mao, T., Oh, J.E., Israelow, B., et al. (2020). Longitudinal analyses reveal immunological misfiring in severe COVID-19. Nature 584, 463–469.

Peng, Y., Mentzer, A.J., Liu, G., Yao, X., Yin, Z., Dong, D., Dejnirattisai, W., Rostron, T., Supasa, P., Liu, C., et al. (2020). Broad and strong memory CD4+ and CD8+ T cells induced by SARS-CoV-2 in UK convalescent individuals following COVID-19. Nat Immunol 1–10.

Richard, J., Prévost, J., Alsahafi, N., Ding, S., and Finzi, A. (2018). Impact of HIV-1 Envelope Conformation on ADCC Responses. Trends Microbiol 26, 253–265.

Risitano, A.M., Mastellos, D.C., Huber-Lang, M., Yancopoulou, D., Garlanda, C., Ciceri, F., and Lambris, J.D. (2020). Complement as a target in COVID-19? Nat Rev Immunol 20, 343–344.

Robbiani, D.F., Gaebler, C., Muecksch, F., Lorenzi, J.C.C., Wang, Z., Cho, A., Agudelo, M., Barnes, C.O., Gazumyan, A., Finkin, S., et al. (2020). Convergent antibody responses to SARS-CoV-2 in convalescent individuals. Nature 584, 437–442.

Rogers, T.F., Zhao, F., Huang, D., Beutler, N., Burns, A., He, W., Limbo, O., Smith, C., Song, G., Woehl, J., et al. (2020). Isolation of potent SARS-CoV-2 neutralizing antibodies and protection from disease in a small animal model. Science 369, 956–963.

Rosado, J., Pelleau, S., Cockram, C., Merkling, S.H., Nekkab, N., Demeret, C., Meola, A., Kerneis, S., Terrier, B., Fafi-Kremer, S., et al. (2020). Serological signatures of SARS-CoV-2 infection: Implications for antibody-based diagnostics. MedRxiv.

Sakurai, A., Sasaki, T., Kato, S., Hayashi, M., Tsuzuki, S., Ishihara, T., Iwata, M., Morise, Z., and Doi, Y. (2020). Natural History of Asymptomatic SARS-CoV-2 Infection. New Engl J Med 383, 885–886.

Schäfer, A., Muecksch, F., Lorenzi, J.C.C., Leist, S.R., Cipolla, M., Bournazos, S., Schmidt, F., Gazumyan, A., Baric, R.S., Robbiani, D.F., et al. (2020). Antibody potency, effector function and combinations in protection from SARS-CoV-2 infection in vivo. Biorxiv 2020.09.15.298067.

Sekine, T., Perez-Potti, A., Rivera-Ballesteros, O., Strålin, K., Gorin, J.-B., Olsson, A., Llewellyn-Lacey, S., Kamal, H., Bogdanovic, G., Muschiol, S., et al. (2020). Robust T cell immunity in convalescent individuals with asymptomatic or mild COVID-19. Cell 183, 158-168.e14.

Varga, Z., Flammer, A.J., Steiger, P., Haberecker, M., Andermatt, R., Zinkernagel, A.S., Mehra, M.R., Schuepbach, R.A., Ruschitzka, F., and Moch, H. (2020). Endothelial cell infection and endotheliitis in COVID-19. Lancet 395, 1417–1418.

Vivier, E., Nunès, J.A., and Vély, F. (2004). Natural Killer Cell Signaling Pathways. Science 306, 1517–1519.

Wang, X., Guo, X., Xin, Q., Pan, Y., Hu, Y., Li, J., Chu, Y., Feng, Y., and Wang, Q. (2020). Neutralizing Antibodies Responses to SARS-CoV-2 in COVID-19 Inpatients and Convalescent Patients. Clin Infect Dis ciaa721-.

Wec, A.Z., Wrapp, D., Herbert, A.S., Maurer, D.P., Haslwanter, D., Sakharkar, M., Jangra, R.K., Dieterle, M.E., Lilov, A., Huang, D., et al. (2020). Broad neutralization of SARS-related viruses by human monoclonal antibodies. Science 369, 731–736.

Wrapp, D., Wang, N., Corbett, K.S., Goldsmith, J.A., Hsieh, C.-L., Abiona, O., Graham, B.S., and McLellan, J.S. (2020). Cryo-EM structure of the 2019-nCoV spike in the prefusion conformation. Science 367, 1260–1263.

Wu, F., Zhao, S., Yu, B., Chen, Y.-M., Wang, W., Song, Z.-G., Hu, Y., Tao, Z.-W., Tian, J.-H., Pei, Y.-Y., et al. (2020). A new coronavirus associated with human respiratory disease in China. Nature 579, 265–269.

Zhang, Q., Bastard, P., Liu, Z., Pen, J.L., Moncada-Velez, M., Chen, J., Ogishi, M., Sabli, I.K.D., Hodeib, S., Korol, C., et al. (2020). Inborn errors of type I IFN immunity in patients with life-threatening COVID-19. Science 370, eabd4570.

Zhou, P., Yang, X.-L., Wang, X.-G., Hu, B., Zhang, L., Zhang, W., Si, H.-R., Zhu, Y., Li, B., Huang, C.-L., et al. (2020a). A pneumonia outbreak associated with a new coronavirus of probable bat origin. Nature 579, 270–273.

Zhou, Z., Ren, L., Zhang, L., Zhong, J., Xiao, Y., Jia, Z., Guo, L., Yang, J., Wang, C., Jiang, S., et al. (2020b). Heightened innate immune responses in the respiratory tract of COVID-19 patients. Cell Host Microbe 27, 883-890.e2.

