## Supplementary figures e for "Asymptomatic and symptomatic SARS-CoV-2 infections elicit polyfunctional antibodies"

Figure S1 (related to Figure 1)

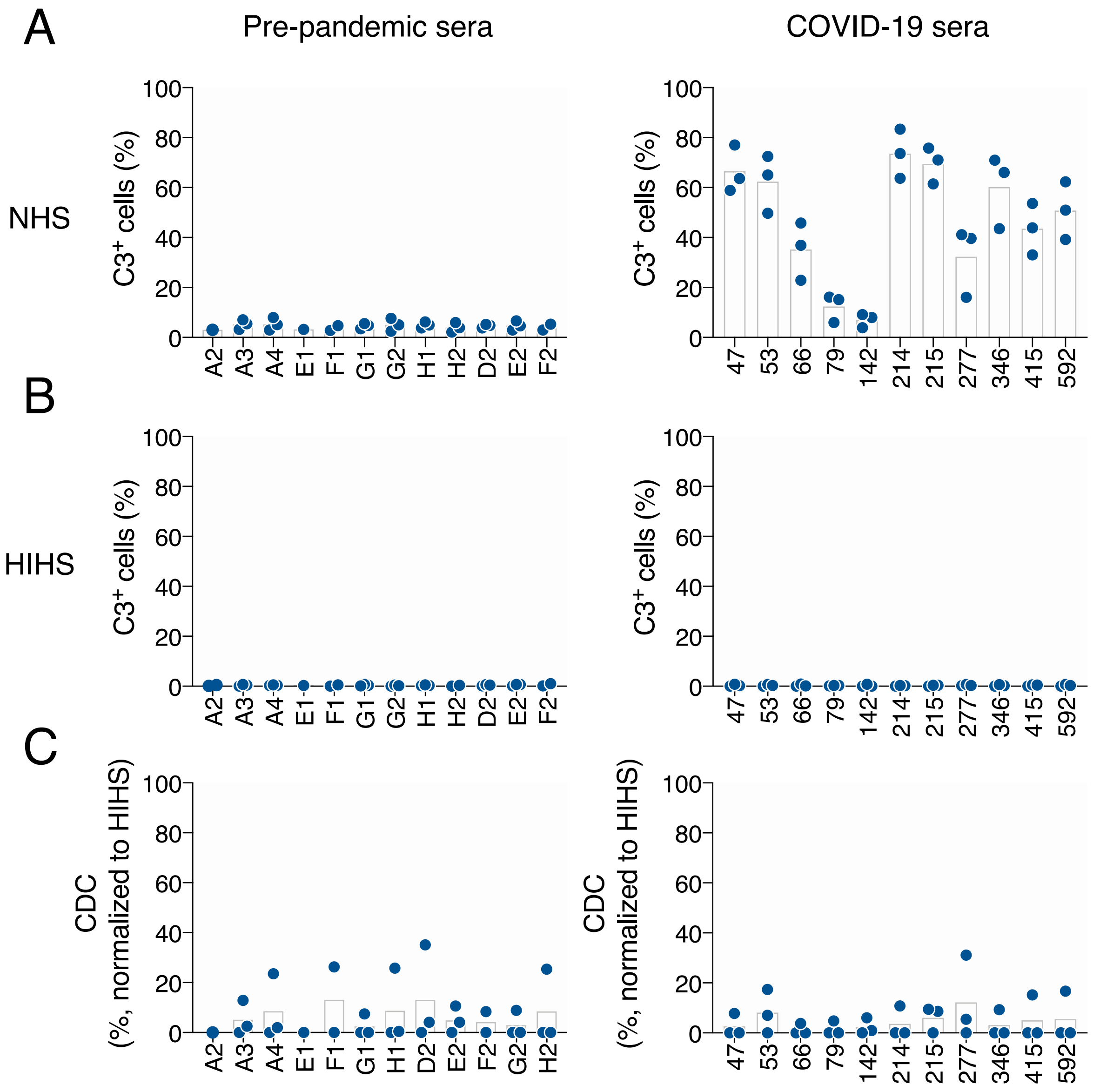

Figure S2 (related to Figure 2)

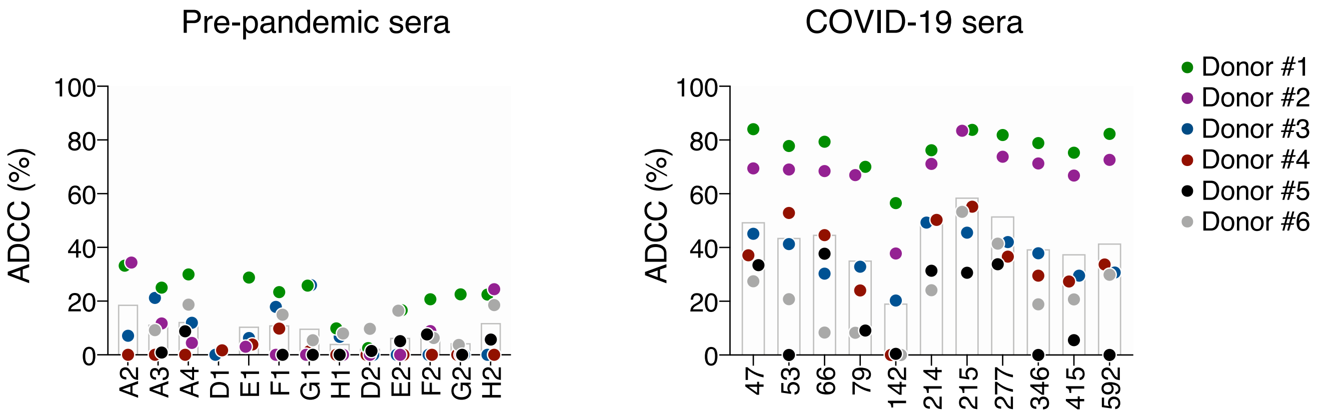

Figure S3 (related to Figure 3)

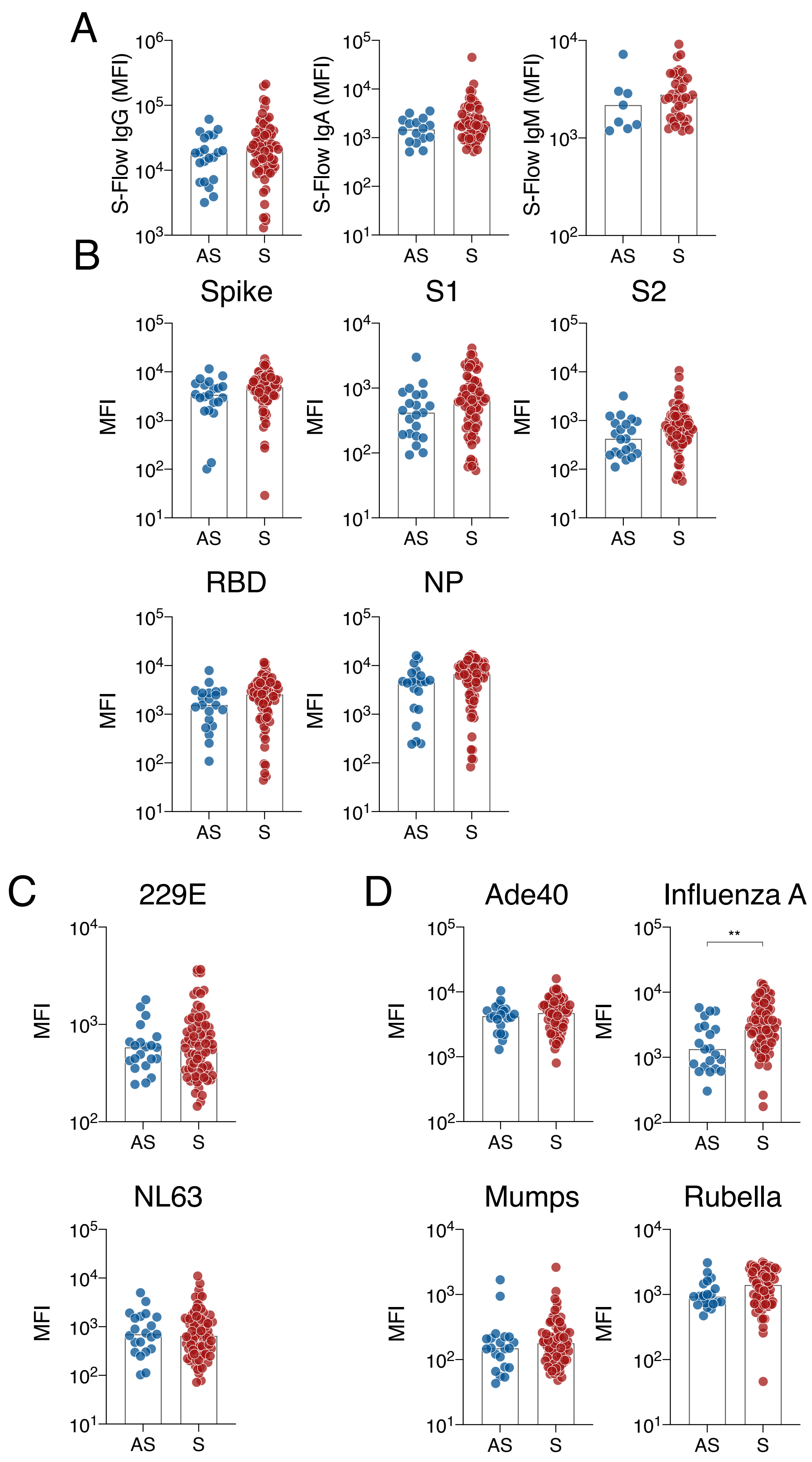

### Figure S4 (related to Figure 5)

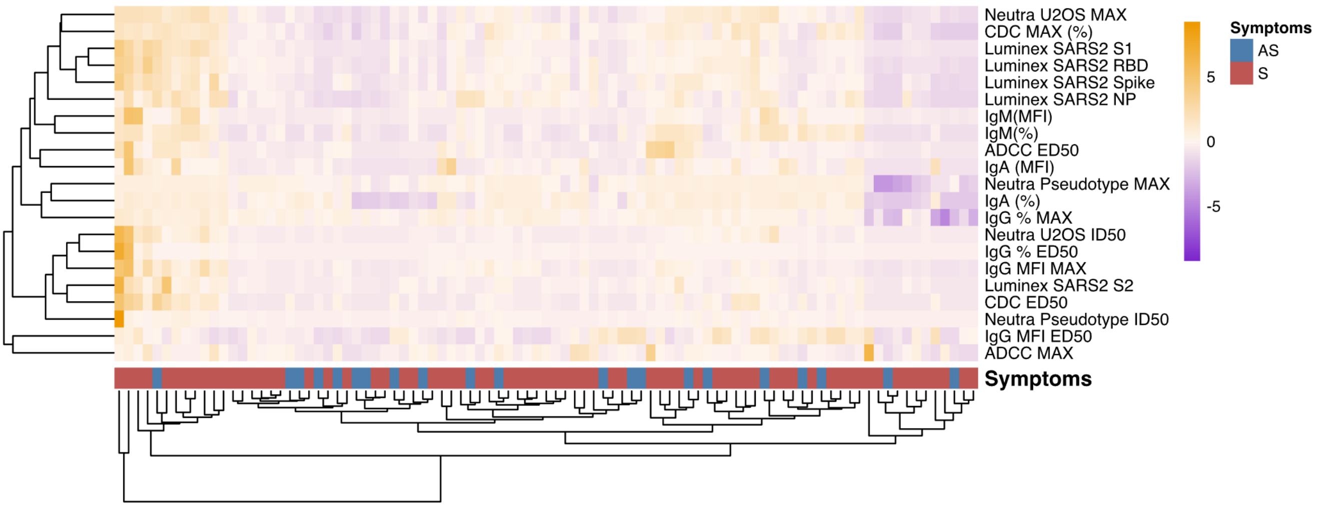

Figure S5 (related to Figure 6 and Table 1 and 2)

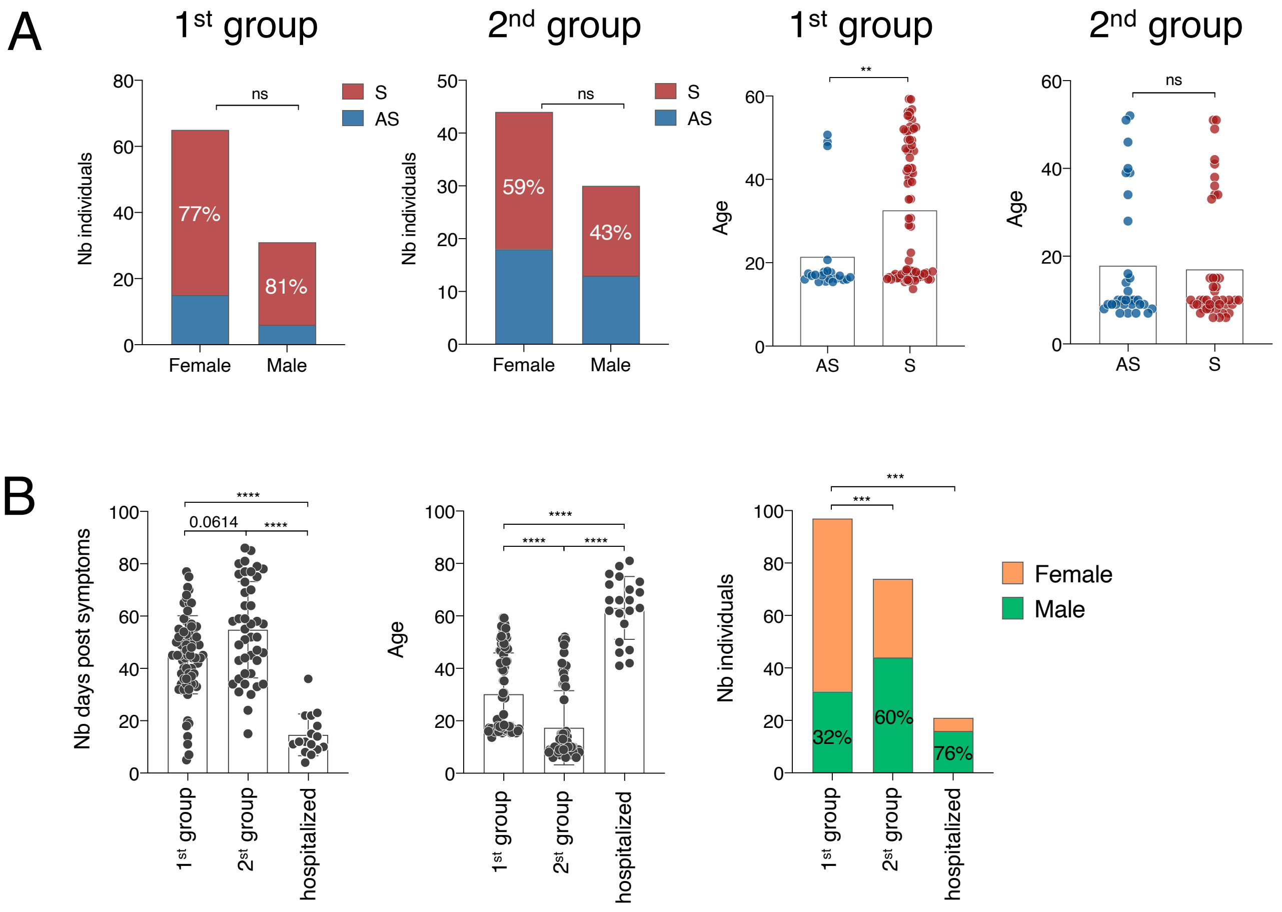

Figure S6 (related to Figure 6)

A

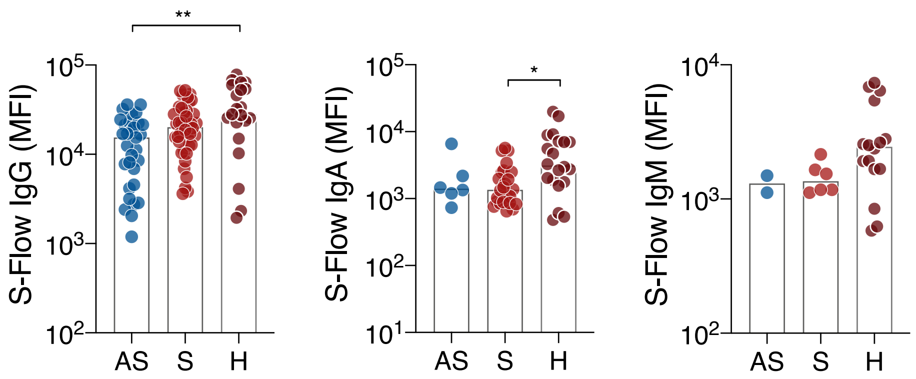


B

| | %, in all individuals | | | MFI, in S-Flow+ individuals | | |
|---|---|---|---|---|---|---|
| Feature | IgG (%) | **IgA (%)** | **IgM (%)** | **IgG (MFI)** | IgA (MFI) | IgM (MFI) |
| AS vs. S p-value (Mann-Whitney test) | ns 0.066 | *** **0.0007** | * **0.0142** | * **0.0177** | ns 0.74 | ns 0.5 |

C

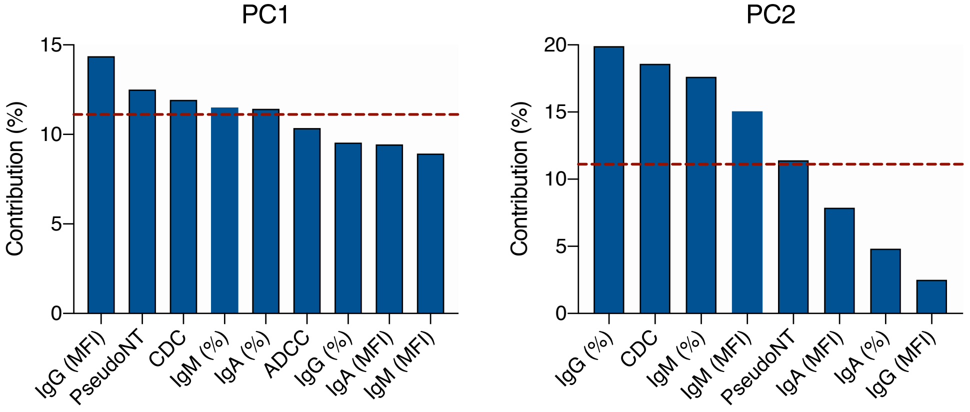

Figure S7 (related to Figure 6)

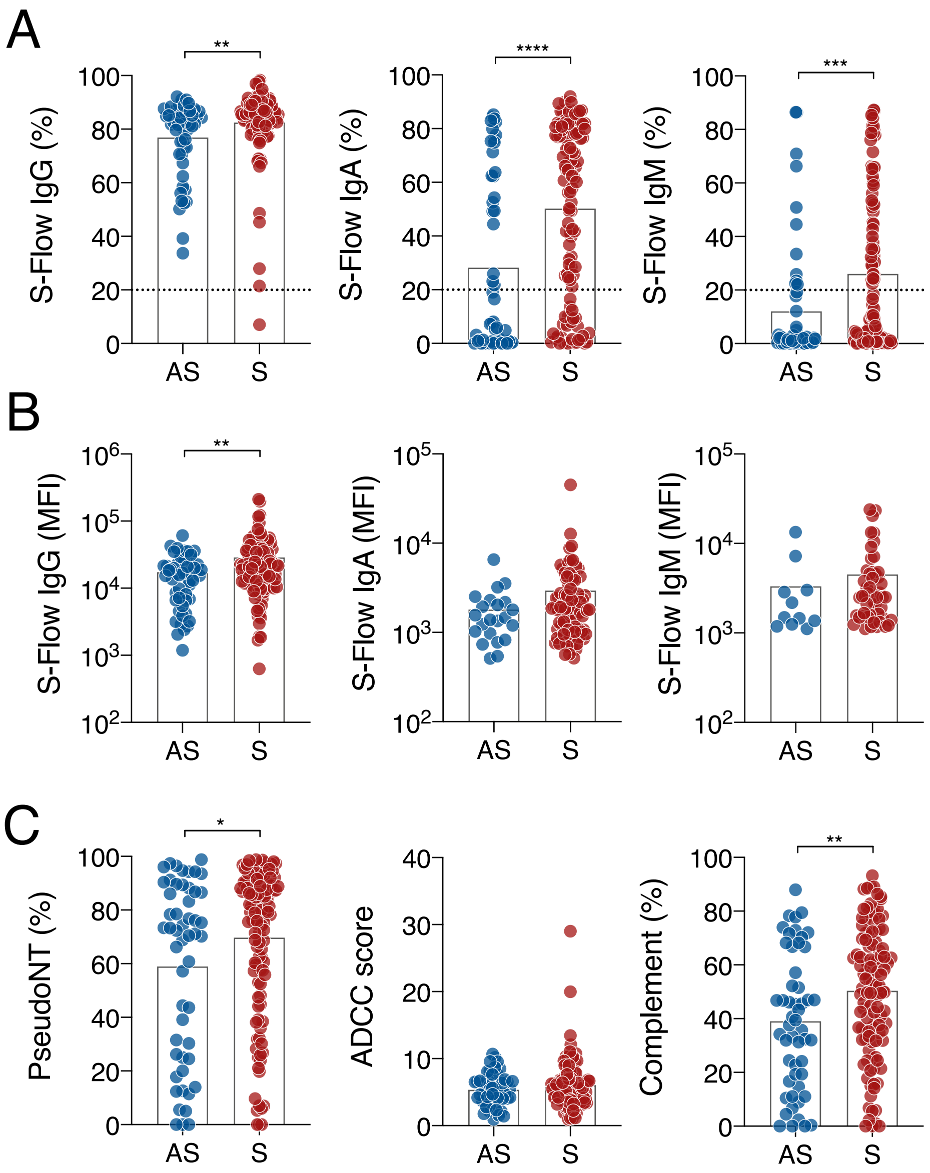
